# From Protocol to Practice: Graded Sepsis Bundle Compliance and Actionable Insights from Real-World ICU Data

**DOI:** 10.64898/2026.04.23.26351412

**Authors:** Himanshu Tripathi, Kaushik Roy, Shahram Rahimi, Subash Neupane, Sean Bozorgzad

## Abstract

Sepsis is a leading cause of in-hospital mortality, yet systematically evaluating temporal adherence to the Surviving Sepsis Campaign (SSC) bundle across large patient populations remains difficult due to semantic variability in electronic health records and the loss of clinical nuance inherent in binary pass/fail compliance judgments. We present an expert-guided neuro-symbolic pipeline that pairs LLM-based semantic normalization with a Sugeno fuzzy inference system encoding eight SSC bundle rules, producing graded per-episode compliance scores whose clinical decision boundaries are set through domain expert consultation. Applied to 2,438 sepsis episodes from MIMIC-IV v3.1, the dual-classifier normalization layer achieves substantial inter-system agreement with high embedding-based confirmation, resolving hundreds of clinically relevant drug strings that purely symbolic systems miss. The graded framework reveals that Hour-1 bundle failures, particularly antibiotic timing, are the dominant driver of low overall compliance, and that higher bundle adherence is associated with notably shorter ICU stays, with antibiotic delays beyond six hours increasing median stays by 61%. These results demonstrate that neuro-symbolic graded assessment can surface actionable compliance patterns that binary evaluation frameworks cannot capture.

**Github:** https://tinyurl.com/yy2zvpd9

## I. Introduction

Sepsis remains a leading cause of in-hospital mortality worldwide, accounting for a significant proportion of deaths in intensive care units and imposing an enormous burden on healthcare systems through prolonged hospitalizations, high treatment costs, and lasting organ damage in survivors. The pathophysiology of sepsis involves a dysregulated host response to infection that can rapidly progress to organ dysfunction, septic shock, and death if not recognized and treated within narrow time windows. Recognizing this urgency, the Surviving Sepsis Campaign (SSC) established evidence-based bundles that specify critical interventions to be completed within the first hour of sepsis recognition, including blood culture collection before antibiotic administration, broad-spectrum antibiotic therapy, and serum lactate measurement, followed by hemodynamic resuscitation and treatment response assessment in subsequent phases [1] (see Appendix: Eight Rules From SSC). Adherence to these bundles has been shown to significantly reduce mortality [2], [3], yet compliance rates remain suboptimal across healthcare institutions globally [4], raising the question of where exactly these failures occur and how they translate into worse patient outcomes. Evaluating temporal adherence to the SSC bundle across large patient populations is a task that carries both clinical urgency and considerable methodological difficulty. Large-scale electronic health record (EHR) databases such as *MIMIC* − *IV* [5] offer an unprecedented opportunity to study sepsis management patterns across thousands of patients and multiple years of clinical practice. However, the clinical data contained in these records is deeply unstructured and inconsistent. Medication names appear as trade names, generic names, abbreviations, and tall-man lettering variants interchangeably, while microbiology results intermix suspected organisms, confirmed pathogens, and contamination events without standardized coding. A clinician documenting `Zosyn` and another documenting `piperacillin-tazobactam` are recording the same antibiotic, yet traditional rule-based compliance systems that rely on exact string matching will fail to recognize one or both of these entries, systematically underestimating true antibiotic administration rates. This semantic variability creates a fundamental barrier to automated compliance assessment that neither purely symbolic systems nor purely neural approaches can overcome alone. Rule-based systems offer transparency and alignment with medical guidelines but break down when confronted with the noise and ambiguity inherent in real-world clinical documentation [6]. Conversely, deep learning models can handle semantic variation and achieve high predictive accuracy, but they operate as black boxes [7] that lack the interpretability and deterministic safety guarantees required for high-stakes clinical decision-making, where the difference between a compliant and non-compliant intervention may depend on whether an antibiotic was given at 58 minutes or 62 minutes after sepsis onset. Beyond the challenge of semantic normalization, existing approaches to compliance assessment suffer from a second critical limitation. Nearly all prior work evaluates bundle adherence using binary judgments, where an intervention either occurred within the guideline window or did not. This binary framing misrepresents clinical reality and discards valuable information about the degree of adherence. A patient receiving antibiotics at 62 minutes is not categorically different from one treated at 58 minutes, yet a binary system assigns them opposite compliance labels. The SSC guidelines themselves acknowledge this nuance by specifying target windows rather than absolute deadlines, recognizing that partial adherence retains clinical value and that the benefit of an intervention decays gradually rather than vanishing at a sharp boundary. What is needed is a graded compliance framework that captures the continuous nature of clinical adherence, assigns proportional credit for nearmiss interventions, and weights different bundle components according to their clinical importance as determined by domain expertise. To address these gaps, we present an expert-guided neuro-symbolic pipeline (Figure 1) that combines LLM-based semantic normalization, domain expert consultation, and a Sugeno fuzzy inference system encoding eight SSC bundle rules to produce graded, temporal compliance scores rather than brittle binary judgments.

**Fig. 1:**
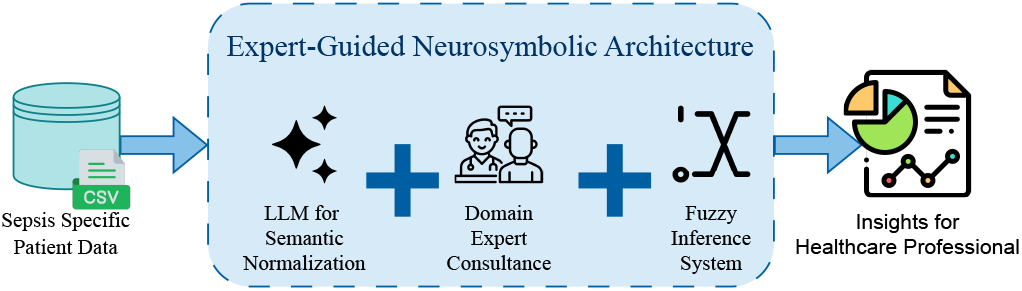
End-to-end workflow of the Expert-Guided Neuro-Symbolic Pipeline, transforming raw sepsis-specific EHR data into actionable clinical insights via three integrated stages: LLM-based semantic normalization of drug and microbiology terminology, domain expert consultation to encode SSC-aligned clinical decision boundaries, and a Sugeno fuzzy inference system evaluating all eight SSC bundle rules to produce graded per-episode compliance scores (0–1) rather than binary pass/fail judgments.

Applied to 2,438 sepsis episodes from *MIMIC* − *IV* v3.1, the pipeline surfaces clinically actionable patterns in bundle adherence and their relationship to patient outcomes. The findings reveal that antibiotic timing is the most critical compliance failure with a mean score of 0.24, that Hour-1 bundle interventions drive systemic underperformance at a mean compliance of just 36.7%, that a striking 51% patient drop-off occurs at the elevated lactate threshold, and that episodes with higher overall bundle compliance achieve notably shorter ICU stays of 3.8 days compared to 5.1 days for low-compliance episodes.

This work is guided by three Research Questions (RQ):

RQ1: Which specific SSC bundle interventions represent the most critical points of clinical failure in sepsis management, and how does a graded compliance framework reveal failure patterns that binary assessment cannot capture?

RQ2: Where do patients fall off the sequential sepsis management pathway, and what do the cascade drop-off patterns reveal about systemic gaps in early sepsis intervention and clinical documentation?

RQ3: How does temporal adherence to SSC bundle components, particularly antibiotic timing and overall compliance, correlate with ICU length of stay and patient outcomes?

The remainder of this paper is organized as follows: Literature Survey reviews related literature, Problem Statement formally defines the core problem, Pipeline Architecture details the pipeline architecture, Quantitative Findings and Expert Discussion presents the results and clinical insights with our final discussion with experts, and lastly, Conclusion concludes the paper. To the best of our ability we have covered all the details within the main paper sections, and please refer to the Appendix for further clarification as needed.

## II. Literature Survey

Sepsis remains a leading cause of mortality in intensive care units, where early detection and timely adherence to evidence-based protocols are critical for patient survival [8]. The Surviving Sepsis Campaign bundles represent the gold standard for reducing mortality [3], yet compliance with these time-sensitive interventions continues to be suboptimal across healthcare settings [4]. Studies have shown that adherence to sepsis bundles significantly impacts patient outcomes [2], making sepsis rules increasingly important. Electronic Health Records such as MIMIC-IV offer unprecedented opportunities for automating sepsis detection and protocol adherence [9], yet these datasets present substantial challenges due to their inherent heterogeneity and complexity. *MIMIC* − *IV* contains a complex mix of high-frequency structured data including vital signs and laboratory results alongside unstructured semantic information such as clinical notes and variable drug names, creating a noisy real-world environment that reflects the true complexity of clinical practice. Purely data-driven approaches such as Deep Learning often achieve high predictive accuracy but operate as black boxes [7], lacking the interpretability and safety guarantees required for high-stakes medical decision-making. Recent studies have demonstrated that machine learning models can identify sepsis early at emergency department triage [10], yet interpretability remains a critical barrier to clinical adoption. Conversely, traditional Rule-Based Systems offer transparency and alignment with medical guidelines but are brittle when facing the noise and ambiguity inherent in real-world clinical data [6], often failing to normalize diverse terminologies or missing context that a human clinician would effortlessly grasp. The challenge is particularly acute in sepsis management where drug names may appear as Vancocin versus Vancomycin, creating barriers for rule-based systems that rely on exact matching and standardized terminology. To effectively address sepsis prediction and protocol adherence in this environment, we need a hybrid system that is both robust to noisy data like Deep Learning [11] models and interpretable regarding safety rules like Expert Systems, enabling reliable automation while maintaining clinical trust and safety. To bridge the gap between the learning capability of neural networks and the reasoning power of symbolic logic, NeSy has emerged as a robust paradigm for healthcare applications [12]. This hybrid approach leverages neural components for perception, handling raw and noisy sensor data, while symbolic components enforce logical constraints and domain knowledge [13]. In the context of medical diagnosis, NeSy architectures allow for the integration of structured biomedical knowledge graphs where neural networks process complex visual or numerical inputs and symbolic reasoning applies domain rules [14]. Recent implementations have shown that combining graph neural networks with symbolic reasoning enables better understanding of biological relationships and enhances clinical decision support systems [15]. In sepsis management specifically, this dual framework becomes valuable as neural models can process time series vital signs while symbolic modules verify compliance with established treatment bundles such as the Surviving Sepsis Campaign guidelines [8]. However, most existing NeSy frameworks rely on standard sequence models like LSTMs or RNNs for their neural component [16]. While these recurrent architectures prove effective for processing time series numerical data such as continuous physiological measurements, they encounter significant limitations when interpreting the semantic nuance embedded in unstructured clinical text [17]. Traditional sequence models struggle to distinguish between subtle but clinically critical textual distinctions such as differentiating a suspected infection from a confirmed diagnosis in free text admission notes [18]. This semantic ambiguity in medical language including negation, uncertainty, and assertion detection presents a fundamental challenge that limits the utility of current NeSy systems in fully automating complex clinical workflows where textual understanding is paramount. Large language models have demonstrated remarkable capabilities in healthcare applications, transforming various aspects of clinical workflow and medical decision support [19]. These models excel at processing complex medical language and extracting meaningful patterns from unstructured clinical narratives [20]. However, despite their linguistic prowess, LLMs exhibit a critical vulnerability that poses substantial risks in clinical environments. Recent investigations have revealed that these models are highly susceptible to adversarial hallucination attacks, where fabricated details embedded in clinical prompts lead to the generation or elaboration of false medical information [21]. This tendency to produce plausible yet factually incorrect outputs represents a fundamental challenge when deploying LLMs in high-stakes medical decision making. Systematic assessment frameworks have been developed specifically to quantify hallucination rates in clinical text summarization, revealing that even state-of-the-art models produce erroneous content at rates that necessitate careful mitigation strategies [22]. The determinism required for critical sepsis interventions, such as fluid resuscitation and antibiotic administration, cannot be guaranteed by purely generative architectures [21], [22]. Recognizing these limitations, researchers have begun exploring hybrid architectures that leverage the semantic understanding capabilities of LLMs while constraining their role to prevent unsafe autonomous decision making. One promising direction involves using LLMs for zero-shot custom feature extraction from clinical notes, where the model functions as an interpretive layer rather than a decision engine [23]. Building on this concept, safety-constrained frameworks have emerged that integrate LLM-based semantic processing with rule-based or symbolic reasoning systems [24]. This architectural pattern aligns well with established fuzzy logic approaches that have proven effective for handling uncertainty in infectious disease diagnosis [25]. Fuzzy logic systems provide a complementary mechanism for uncertainty-aware decision support when processing heterogeneous medical data [26]. The integration of fuzzy logic with LLM-based semantic normalization has also been explored in systematic review pipelines, where LLMs interpret complex medical language while fuzzy logic handles the classification and screening decisions [27]. This emerging paradigm positions the LLM as a Semantic Normalizer that translates the messy heterogeneity of electronic health record data into structured inputs, while preserving the deterministic safety guarantees provided by symbolic reasoning engines for actual clinical decision making. The overall expert-guided neuro-symbolic workflow is illustrated in Figure 2. Such an architecture directly addresses the dual challenge of handling linguistic complexity while maintaining clinical safety standards in sepsis management protocols.

**Fig. 2:**
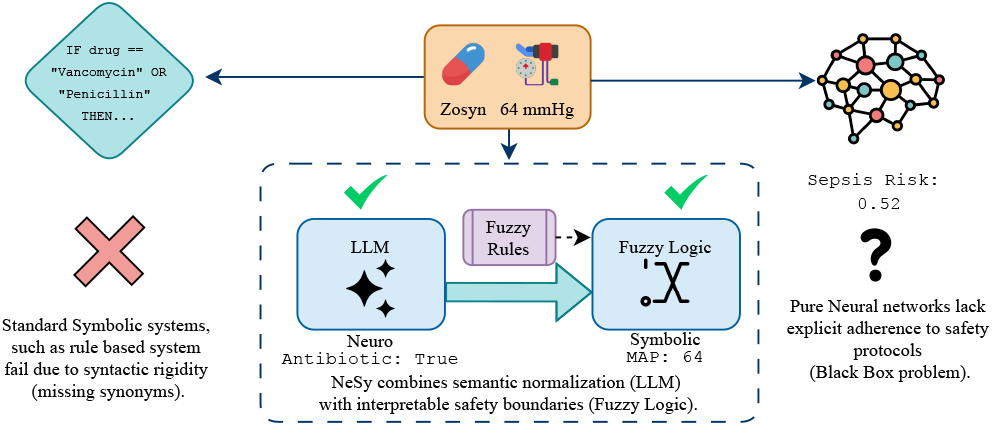
Comparison of three sepsis management paradigms applied to the antibiotic trade name `Zosyn` and a MAP of 64 mmHg. Rule-based symbolic systems fail from syntactic rigidity, misclassifying `Zosyn` and systematically underestimating antibiotic compliance. Pure neural networks resolve semantic variation but produce opaque risk estimates incompatible with the determinism required for high-stakes clinical decisions. The proposed Neuro-Symbolic (NeSy) architecture resolves both limitations by pairing LLM semantic normalization with deterministic fuzzy logic safety boundaries.

## III. Problem Statement

The guidelines from the Surviving Sepsis Campaign demand that key interventions such as administering antibiotics and fluids, measuring lactate levels, and obtaining blood cultures, be executed within strict time limits. Assessing adherence to these protocols on a massive scale is currently impractical. This difficulty arises because medical records freely mix generic, brand, and abbreviated drug names, while microbiology reports blend actual pathogens with suspected organisms and mere contaminants, erecting significant hurdles for automated evaluation. Conventional rule-based approaches cannot handle this intricate variability. For instance, if a provider writes `Zosyn`, a strict rule designed to find `piperacillin-tazobactam` will miss it because of its inflexible matching criteria. Here, *v* represents the specific variant found in the record, and *V* represents the pre-established list of synonyms:

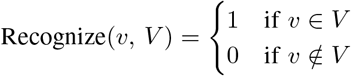

**TABLE I:** Notation and Symbols for Problem Statement Formulation.

| Symbol | Definition |
| --- | --- |
| $v$ | Documented drug variant in clinical records |
| $V$ | Predefined synonym set in rule-based system |
| $\mathcal{L}$ | Large Language Model for semantic normalization |
| $\text{input}_m$ | Clinical prompt supplied to $\mathcal{L}$ |
| $S_k$ | Sepsis episode $k$ |
| $\mathbb{P}$ | $\mathbb{P}(E)$ denotes probability of event $E$ |
| $\mathcal{D}$ | Clinical decision boundary (expert-validated threshold) |
| $\mathcal{F}$ | Fuzzy Inference System encoding SSC logic |

While Large Language Models ℒ can untangle this linguistic diversity, they carry an unmeasured risk of hallucination. Specifically, the likelihood ℙ that the neural model’s outputs are firmly anchored in actual documented evidence remains unknown:

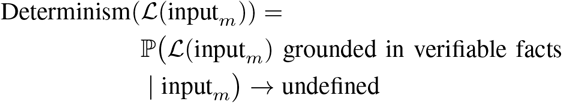

In this context, input_*m*_ stands for the specific clinical prompt given to ℒ. Currently, no framework generates a tiered evaluation of protocol adherence by merging language standardization with clear, rule-based logic. To solve this, we introduce an Expert-Guided Neuro-Symbolic Pipeline. This system restricts ℒ entirely to the task of standardizing terminology, deliberately blocking it from making any independent clinical judgments. Subsequently, a Fuzzy Inference System ℱ evaluates the sepsis case *S*_*k*_ using decision thresholds confirmed by medical experts 𝒟 to generate scaled compliance metrics:

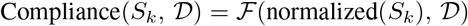

These resulting metrics provide practical, data-driven insights for healthcare professionals, demonstrated across 2,438 distinct sepsis episodes drawn from the *MIMIC* − *IV* dataset.

## IV. Pipeline Architecture

The Expert-Guided Neuro-Symbolic Pipeline (Figure 3) uses the hybrid approach outlined in the Problem Statement by integrating semantic normalization, expert validation, and fuzzy reasoning into a cohesive workflow. This section details the complete architecture beginning with data preparation from *MIMIC* − *IV* . We first describe the preprocessing steps in Cohort Selection and Data Preparation, followed by the semantic normalization component in Semantic Normalization where the LLM processes unstructured clinical text. The validation framework ensuring semantic grounding is discussed in Validation, while Domain Expert Consultation outlines the expert consultation process for establishing clinical decision boundaries. Finally, Fuzzy Compliance Assessment describes the Fuzzy Inference System that transforms normalized features into graded compliance assessments. Refer to Appendix:Data Overview for the overall *MIMIC* − *IV* data landscape.

**Fig. 3:**
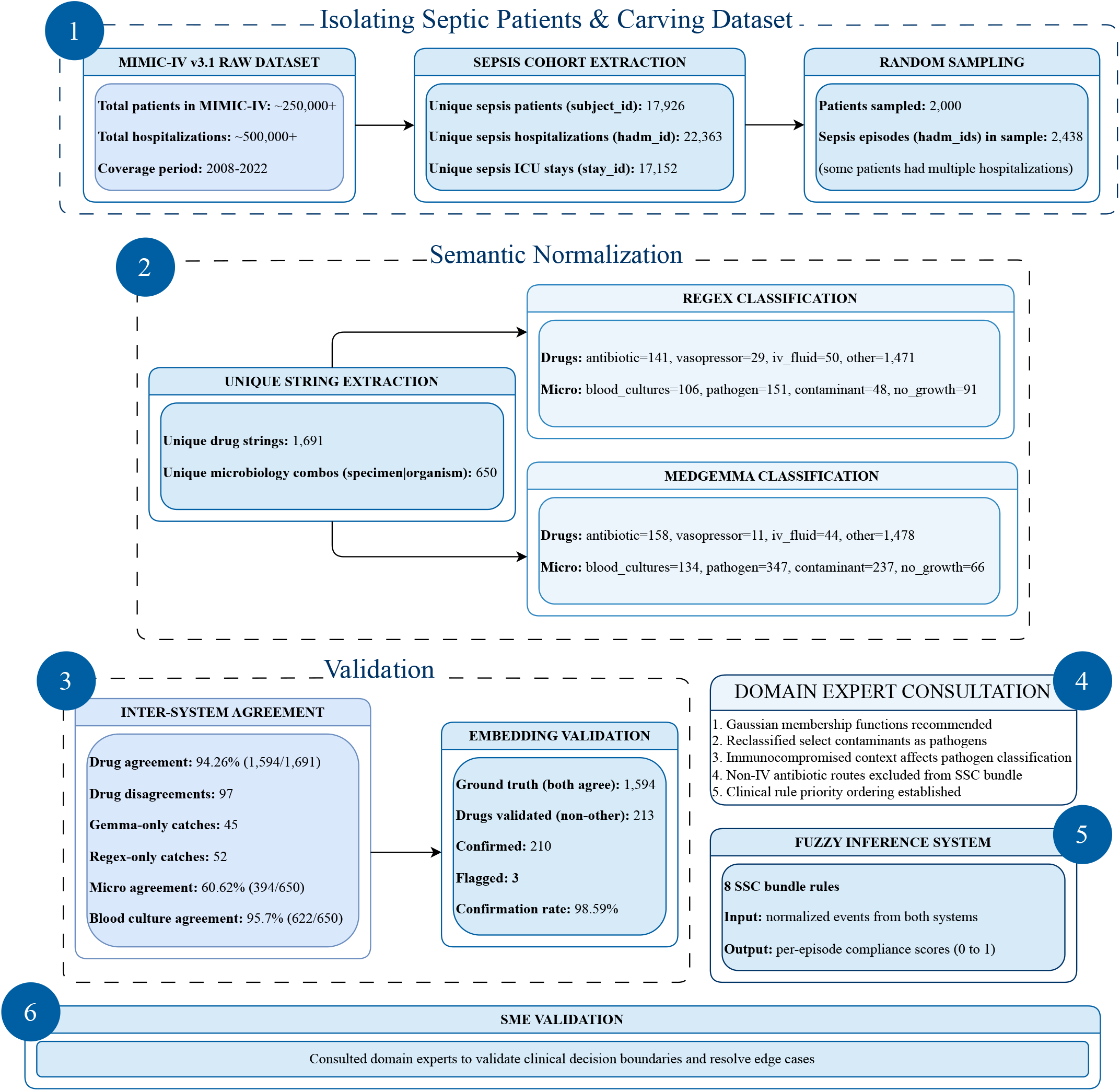
detailed pipeline architecture across six stages applied to 2,438 sepsis episodes from mimic-iv v3.1: (1) icd-9/icd-10 sepsis cohort extraction; (2) dual regex and medgemma-4b-it classification of 1,691 drug strings and 650 microbiology combinations; (3) inter-classifier validation (94.26% drug agreement, *κ*=0.65) confirmed by embedding-based cosine similarity (98.59% confirmation rate); (4) domain expert consultation establishing gaussian membership functions, immunocompromised-host pathogen reclassification, iv/im-only antibiotic criteria, and rule priority ordering; (5) sugeno fuzzy inference across eight ssc rules; and (6) sme edge-case resolution.

### A. Cohort Selection and Data Preparation

Sepsis episodes were identified from *MIMIC* − *IV v*3.1 using *ICD* − 10 codes *A*40, *A*41, *R*65.20, and *R*65.21 alongside *ICD* − 9 codes 038.*, 995.91, 995.92, and 785.52, searched in both coding versions because *MIMIC* − *IV* spans the 2008-2022 transition between *ICD* eras and restricting to a single era would systematically exclude a substantial portion of the sepsis population. This identified 17,926 unique patients across 22,363 hospitalizations with 17,152 ICU stays (Figure 3 (1)). Random sampling with seed 55 produced a cohort of 2,000 patients and 2,438 sepsis episodes, with some patients contributing multiple hospitalizations, a clinically meaningful feature reflecting recurrent sepsis associated with immunosuppression and chronic comorbidities that enables analysis of whether compliance improves across repeated encounters. The cohort exceeds the minimum sample sizes required by power analysis for Cohen’s Kappa (*n* = 22, exceeded 76.9×), McNemar’s test (*n* = 471, exceeded 5.2×), and fuzzy membership function estimation (*n* = 240, exceeded 10.2 ×).

Data preparation transforms the raw relational structure into an episode-centric format. For each hospitalization, the pipeline creates seven structured files: `medications_admin.csv` (eMAR timestamps), `medications_ordered.csv` (prescriptions), `microbiology.csv` (culture specimens and organisms), `labs.csv` (lactate and laboratory values), `vitals.csv` (blood pressure, heart rate, temperature), `inputs.csv` (IV fluids and vasopressor infusion rates), and `static_profile.json` (demographics, ICU timestamps, body weight). Each file maintains minute-level temporal resolution enabling precise alignment with SSC protocol windows. This structure directly mirrors bundle information requirements: blood culture sequencing (R1) requires cross-referencing microbiology against medication times, antibiotic (R2) and lactate timing (R3) require anchoring to sepsis onset, fluid adequacy (R5) demands body weight for the 30 mL/kg target, and vasopressor assessment (R6, R7) requires linking infusion data with concurrent vital signs.

### B. Semantic Normalization

Clinical documentation embeds substantial semantic noise that threatens compliance measurement accuracy. Clinicians record antibiotics (Listing 1) as trade names (`Zosyn`, `Rocephin`), generic names (`piperacillin-tazobactam`), abbreviations (`CTX`), and tall-man lettering (`DAPTOmycin`), while microbiology results (Listing 2) vary between “blood culture”, “BC”, and “blood cx” without consistent coding. Failing to recognize `Zosyn` as an antibiotic means a patient who received timely therapy would be incorrectly scored as non-compliant. The normalization strategy extracts unique strings once, classifies them centrally, and maps labels back across all episodes, identifying 1,691 unique drug strings and 650 microbiology combinations (Figure 3(2)).

**Listing 1:**
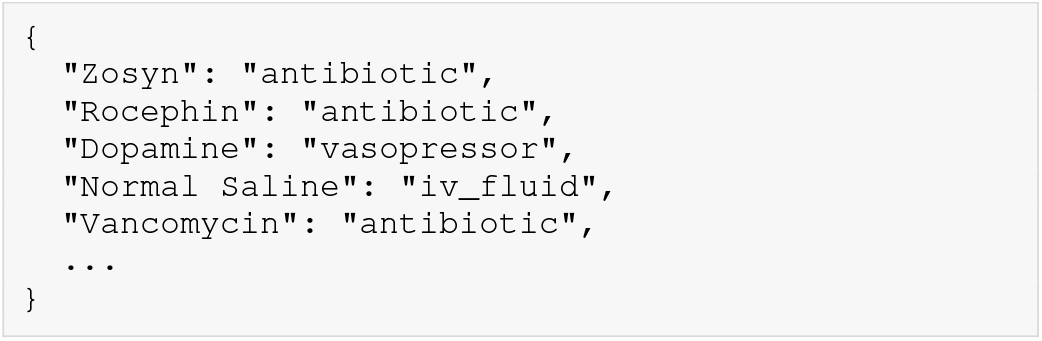
Drug Classification: 5 classified examples from 1,594 total agreements

Two complementary classifiers are applied. First, regex with domain-informed exclusion rules classifies drugs into antibiotic, vasopressor, IV fluid, and other, with clinically motivated exclusions: topical formulations filtered because SSC mandates IV/IM administration, saline flushes excluded as catheter maintenance, and hypertonic saline excluded as osmotic therapy. Microbiology classification identifies blood cultures, then categorizes organisms as pathogen, contaminant (skin flora in immunocompetent patients), or no growth. Regex yielded 141 antibiotics, 29 vasopressors, 50 IV fluids, and 1,471 other, with 106 blood culture specimens, 511 pathogens, 48 contaminants, and 91 no-growth. Second, MedGemma-4b-it under zero-shot structured prompting produced 158 antibiotics, 11 vasopressors, 44 IV fluids, and 1,478 other, with 134 blood culture specimens, 347 pathogens, 237 contaminants, and 66 no-growth. The microbiology divergence reflects MedGemma incorporating specimen-site context more aggressively, mirroring how infectious disease specialists interpret culture results where pathogen significance depends on clinical context. Both classifiers are necessary because their error profiles are complementary: regex achieves near-perfect precision on explicit matches but zero recall on synonyms, while MedGemma generalizes across variations at the cost of occasional hallucination. Drug agreement reached 94.26% and microbiology agreement 60.62%.

### C. Validation

Agreement alone does not guarantee correctness, as two classifiers can converge on the same wrong answer, and in a clinical context where misclassifying a vasopressor as an antibiotic would distort compliance scores with implications for quality metrics and patient safety assessments, establishing validity is essential. Drug agreement reaches 94.26% (1,594*/*1,691), with 97 disagreements splitting into 45 Gemma-only and 52 regex-only catches, revealing that neither classifier is systematically superior. Microbiology agreement is 60.62% (394*/*650), reflecting different classification philosophies, while blood culture detection, the clinically critical question anchoring Rule 1, achieves 95.7% agreement (622*/*650). Cohen’s Kappa *κ* = (*P*_*o*_ − *P*_*e*_)*/*(1 − *P*_*e*_) with *P*_*o*_ = 0.9426 and *P*_*e*_ = 0.8277 (from the confusion matrix, −Figure F) yields *κ* = 0.65 (substantial agreement). McNemar’s test *χ*^2^ = (*b* −*c*)^2^*/*(*b* + *c*) = (45 − 52)^2^*/*97 = 0.505 produces *p* = 0.088, failing to reject marginal homogeneity and confirming complementary rather than redundant error profiles, a property essential for a clinical pipeline where directional bias could systematically inflate or deflate scores for specific intervention types.

**Listing 2:**
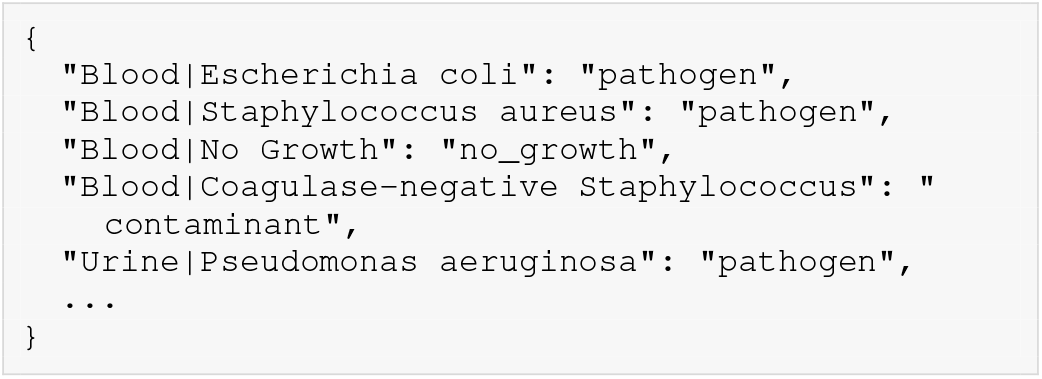
Microbiology Classification: 5 classified examples from 394 total agreements (`“Blood | Coagulase-negative Staphylococcus”: “contaminant”` is for immunocompetent patient)

To verify that agreement reflects genuine semantic understanding rather than shared systematic error, we exploit MedGemma’s embedding space as an independent reference. The 1,594 agreed classifications serve as ground truth to compute per-category cosine similarity thresholds (*θ*_antibiotic_ = 0.3206, *θ*_vasopressor_ = 0.2263, *θ*_iv fluid_ = 0.3075) using sim(*d*_*i*_, *a*_*k*_) = **e**(*d*_*i*_) · **e**(*a*_*k*_)*/*(∥**e**(*d*_*i*_) ∥ ∥ **e**(*a*_*k*_) ∥), where **e**(·) denotes the mean-pooled token embedding. All 213 non-`other` Gemma classifications are tested: 210 confirmed, 3 flagged due to ALL-CAPS tall-man lettering tokenizer artifacts (e.g., `DAPTOmycin`), yielding a 98.59% confirmation rate (Figure 3(3)). The 97 disagreements are adjudicated by comparing each drug’s embedding against both claimed category anchors, yielding 49 regex wins and 48 Gemma wins, a near-symmetric split independently confirming the McNemar result. The clinical consequence is that 220 drug strings that would have been missed or misclassified by either classifier alone are correctly resolved, ensuring semantically accurate inputs for the downstream compliance engine (Figure 3(4)).

### D. Domain Expert Consultation

Subject matter experts were integrated mid-pipeline rather than retrospectively, ensuring clinical knowledge shaped both normalization rules and fuzzy parameters before any compliance scores were generated (Figure 3 (6)). Domain clinicians reviewed classifier outputs alongside the complete pipeline architecture and provided four critical inputs. First, they recommended Gaussian membership functions over trapezoidal or triangular alternatives, reasoning that clinical benefit does not suddenly vanish at a boundary but degrades gradually as delays accumulate, and that a patient receiving antibiotics at 65 minutes has not lost all therapeutic value compared to one treated at 59 minutes. Second, experts reclassified organisms based on immunocompromised status, establishing that coagulase-negative staphylococci, typically skin contaminants in immunocompetent patients, should be treated as true pathogens in immunocompromised hosts where they cause genuine bloodstream infections, directly affecting Rule 1 scoring by determining whether a positive culture represents a meaningful diagnostic event. Third, clinicians mandated route-specific exclusion rules: only IV/IM antibiotic administration counts toward SSC compliance, excluding topical preparations, oral prophylaxis, and ophthalmic formulations that do not achieve systemic therapeutic concentrations relevant to sepsis management. Fourth, experts established the rule priority ordering *R*_3_ *> R*_2_ *> R*_5_ *> R*_6_ *> R*_1_ = *R*_4_ = *R*_7_ = *R*_8_, reflecting clinical consensus that lactate measurement and antibiotic administration are the most time-sensitive and outcome-determining interventions and should exert proportionally greater influence on overall compliance scores than downstream response metrics. These four decisions collectively shaped the normalization exclusion patterns, the membership function parameterization, the microbiology interpretation logic, and the weighted defuzzification formula, ensuring that every computational decision in the pipeline is grounded in expert-validated clinical reasoning rather than arbitrary algorithmic choices.

### E. Fuzzy Compliance Assessment

Binary compliance evaluation misrepresents clinical reality: a patient receiving antibiotics at 62 minutes is not categorically different from one treated at 58 minutes. Following Domain Expert Consultation, the pipeline employs a Sugeno fuzzy inference system producing graded scores via half-Gaussian membership functions. For timing-based rules, *μ*_right_(*x*; *c, σ*) = 1 if *x* ≤ *c*, else exp(− (*x* − *c*)^2^*/*2*σ*^2^). For value-based rules, *μ*_left_(*x*; *c, σ*) = exp(− (*x* −*c*)^2^*/*2*σ*^2^) if *x < c*, else 1. A window variant *μ*_window_ penalizes both premature and delayed action (Figure 4).

**Fig. 4:**
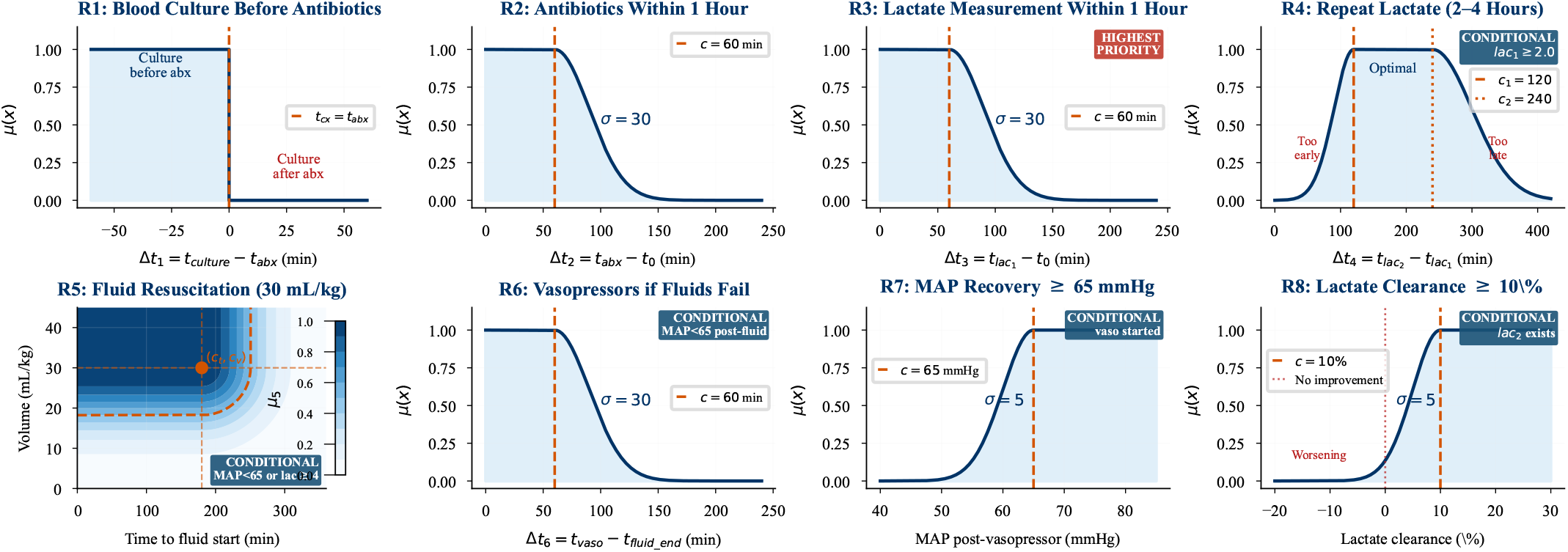
Fuzzy membership functions *μ*(*x*) for all eight SSC bundle rules, with clinical thresholds established through domain expert consultation. Phase 1 rules encode Hour-1 mandates: R1 (Boolean, culture before antibiotics), R2 and R3 (right-sided half-Gaussian, *c*=60 min, *σ*=30), and R4 (window function, optimal repeat lactate at 2–4 hours, conditional on lactate *≥* 2.0 mmol/L). Phase 2 rules encode hemodynamic resuscitation: R5 (2D contour combining fluid timing and 30 mL/kg volume target) and R6 (vasopressor timing, *c*=60 min). Phase 3 rules encode physiologic recovery targets: R7 (MAP ≥ 65 mmHg, *σ*=5) and R8 (lactate clearance ≥ 10%, *σ*=5). Purple badges denote conditional rules that activate only when specified hemodynamic or metabolic triggers are met.

#### Phase 1 (Hour-1 Bundle)

R1 is Boolean (*z*_1_ = 1 if *t*_culture_ *< t*_abx_, else 0), ensuring cultures precede antibiotics to preserve pathogen identification. R2 evaluates antibiotic timing via *μ*_right_(Δ*t*_2_; 60, 30), where treatment at 90 min yields *μ* = 0.61 and at 120 min yields *μ* = 0.13, capturing the dose-response relationship between delay and mortality. R3 (highest priority) applies the same parameterization to lactate measurement, foundational to all subsequent resuscitation decisions. R4 activates when lactate ≥ 2.0 mmol/L, evaluating repeat measurement via *μ*_window_(Δ*t*_4_; 120, 240, 30, 60) with asymmetric decay (*σ*_early_ = 30, *σ*_late_ = 60) reflecting that premature re-testing provides less useful information.

#### Phase 2 (Resuscitation)

R5 activates when MAP *<* 65 mmHg or lactate ≥ 4.0 mmol/L, combining temporal (*μ*_right_(Δ*t*_5_; 180, 60)) and volumetric (*μ*_left_(*V/W* ; 30, 10)) components via *z*_5_ = *μ*_5*t*_ × *μ*_5*v*_. R6 evaluates vasopressor timeliness via *μ*_right_(Δ*t*_6_; 60, 30) when MAP remains below 65 mmHg post-fluids.

#### Phase 3 (Response)

R7 assesses MAP recovery via *μ*_left_(MAP; 65, 5), where *σ* = 5 reflects the narrow margin before ischemic organ damage. R8 evaluates lactate clearance via *μ*_left_(clearance; 10, 5), with negative values producing scores near zero.

Missing data handling follows SSC philosophy: untriggered conditional rules receive *μ* = 1.0 and are excluded from *R*_*i*_, mandatory rules (R1-R3) with missing data receive *μ* = 0 since undocumented care cannot be credited, and rules with unevaluable triggers are excluded entirely. Per-episode compliance aggregates via weighted Sugeno defuzzification: Compliance 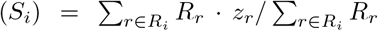, where *R*_*r*_ encodes the expert priority ordering. Each episode is evaluated twice (regex and MedGemma paths), producing identical scores where classifiers agree (94.26%) and quantifying clinical impact of classification differences where they diverge.

### V. Quantitative Findings with Expert Discussion

The application of our expert-guided neuro-symbolic pipeline to 2,438 sepsis episodes from the MIMIC-IV v3.1 database reveals several clinically important patterns in how hospitals adhere to the Surviving Sepsis Campaign bundle. Before examining compliance itself, it is essential to establish the reliability of the semantic normalization layer that feeds the fuzzy inference system. The regex and MedGemma classifiers achieved 94.26% agreement across 1,691 unique drug strings, with a Cohen’s kappa of 0.65 indicating substantial concordance. Of the 2,438 episodes evaluated, the vast majority produced identical compliance scores under both classifiers, and only 80 episodes (3.3%) showed any divergence at all, with just 53 of those exceeding a 5% score difference (Figure 5). This tight agreement confirms that the downstream compliance scores are robust to the choice of classifier and that the small number of adjudicated disagreements does not materially alter population-level findings. Importantly, the semantic normalization component resolved 220 clinically relevant drug strings that a purely pattern-based system would have missed, including brand name variants such as `Zosyn` for piperacillintazobactam and combination formulations that do not follow standard naming conventions. Turning to the clinical findings, the eight-rule compliance profile (Figure 6) exposes a stark and medically concerning stratification. Antibiotic timing, encoded as Rule 2, emerges as the single most critical compliance failure with a mean fuzzy score of just 0.24, placing it firmly in the critical performance zone. Only 13% of episodes achieved antibiotic administration within the strict one-hour target mandated by the SSC guidelines, a figure that initially appears alarming but requires clinical contextualization. Discussion with domain experts revealed a plausible explanation for this finding. By the time a septic patient is formally admitted to the intensive care unit, clinicians typically already suspect sepsis, and antibiotics are frequently administered in the emergency department or on the ward prior to ICU transfer. The observed timing distribution (Figure 12) corroborates this interpretation, showing a substantial proportion of episodes where antibiotic administration preceded the sepsis onset timestamp recorded in the MIMIC-IV database. Because the pipeline anchors all timing measurements to this recorded onset, pre-ICU antibiotic administration is penalized even when it may represent appropriate early clinical action. This finding highlights a fundamental tension between retrospective database analysis and real-world clinical workflows, where the moment of clinical suspicion rarely aligns perfectly with the documented timestamp of formal sepsis recognition.

**Fig. 5:**
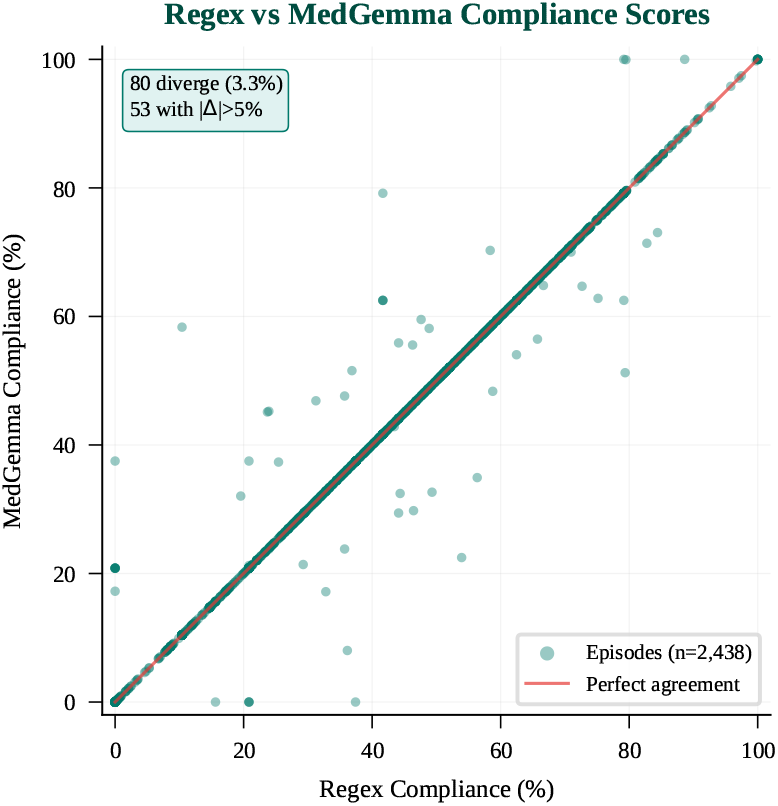
Inter-classifier concordance scatter plot comparing SSC bundle compliance scores derived from regex and MedGemma normalizers across 2,438 sepsis episodes. Only 80 episodes (3.3%) show any score divergence, with 53 (2.2%) exceeding a 5% difference, confirming that population-level compliance findings are robust to classifier choice and that the dual-classifier consensus does not materially distort cohort-level adherence estimates.

**Fig. 6:**
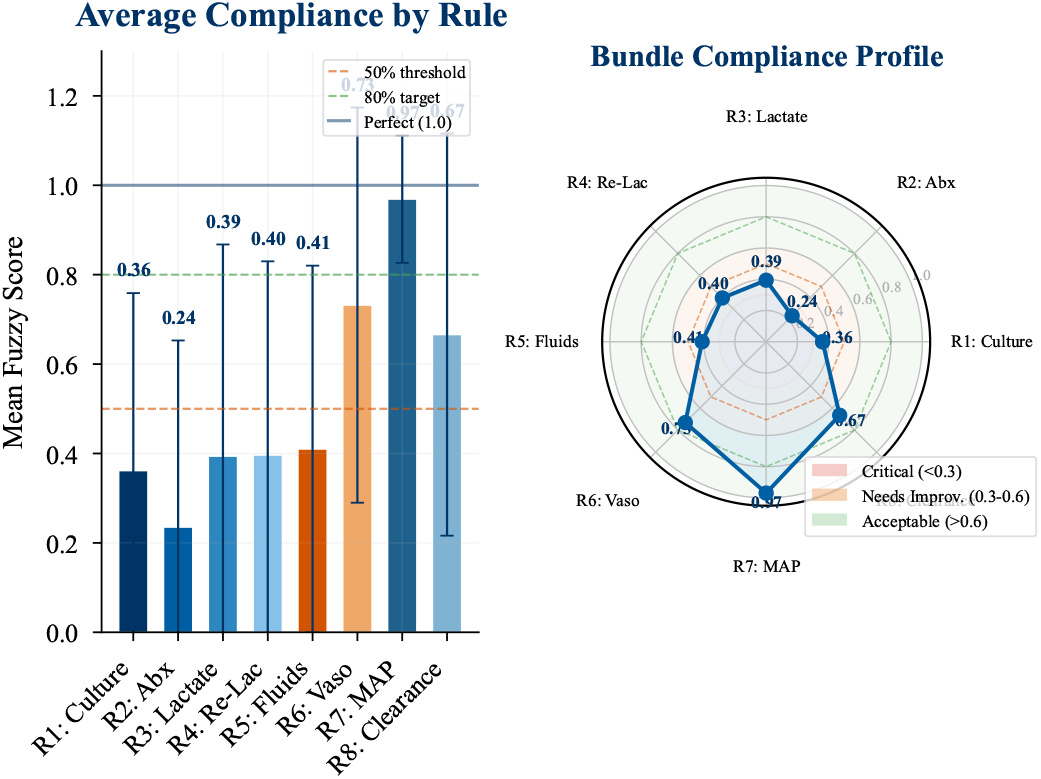
**Left:** Mean fuzzy compliance scores for all eight SSC bundle rules, with reference lines at the 50% minimum threshold, 80% quality target, and perfect adherence (1.0). Antibiotic timing (R2, *μ*=0.24) is the sole rule in the critical zone (*<*0.3); blood culture sequencing (R1, *μ*=0.36) and initial lactate measurement (R3, *μ*=0.39) also fall below 50%, collectively driving Hour-1 underperformance. High vasopressor (R6, *μ*=0.73) and MAP recovery (R7, *μ*=0.97) scores reflect survivorship bias rather than genuine clinical excellence. **Right:** Radar chart displaying the full bundle compliance profile stratified by performance tier: critical (*<*0.3), needs improvement (0.3–0.6), and acceptable (*>*0.6).

Blood culture sequencing (R1, mean score 0.36) and initial lactate measurement (R3, mean score 0.39) also fall below the 50% compliance threshold, indicating systemic failures in the earliest phase of sepsis management. These three rules together form the core of the Hour-1 bundle, and their collective underperformance drives the overall compliance distribution, which shows a mean of 36.7% and a median of 37.5% across all episodes (Figure 11). By contrast, the conditional hemodynamic rules tell a markedly different story. Vasopressor initiation (R6, mean 0.73) and mean arterial pressure recovery (R7, mean 0.97) appear near acceptable or even exemplary. However, this apparent high performance likely reflects a survivorship bias artifact rather than genuine clinical excellence. These conditional rules only activate in patients who have survived long enough to reach the secondary phase of care, where shock has been recognized and initial fluid resuscitation has proven insufficient. At this stage, clinical protocols for vasopressor escalation are well established and consistently executed, creating the misleading impression that downstream interventions are performed reliably while masking the upstream failures that determine whether patients ever reach this treatment threshold.

The compliance cascade analysis (Figure 7) provides a sequential view of where patients fall off the bundle pathway and reveals the most alarming finding of this study. Beginning with all 2,438 sepsis episodes, 77% had blood cultures obtained, and 68% received an IV or intramuscular antibiotic, representing moderate but not catastrophic initial drop-offs. Lactate was measured in 76% of episodes, a reasonably high proportion. However, a striking 51% drop occurs at the elevated lactate threshold of 2.0 mmol/L or higher, with only 912 episodes (37%) recording this value. Domain experts flagged this as particularly concerning because it indicates that the majority of sepsis episodes either had lactate levels that were never elevated, were never measured at all, or were measured but not documented. The missing data analysis (Figure 14) supports this concern, revealing that antibiotic timing (31.6%) and lactate measurement (24.4%) are the most data-sparse

**Fig. 7:**
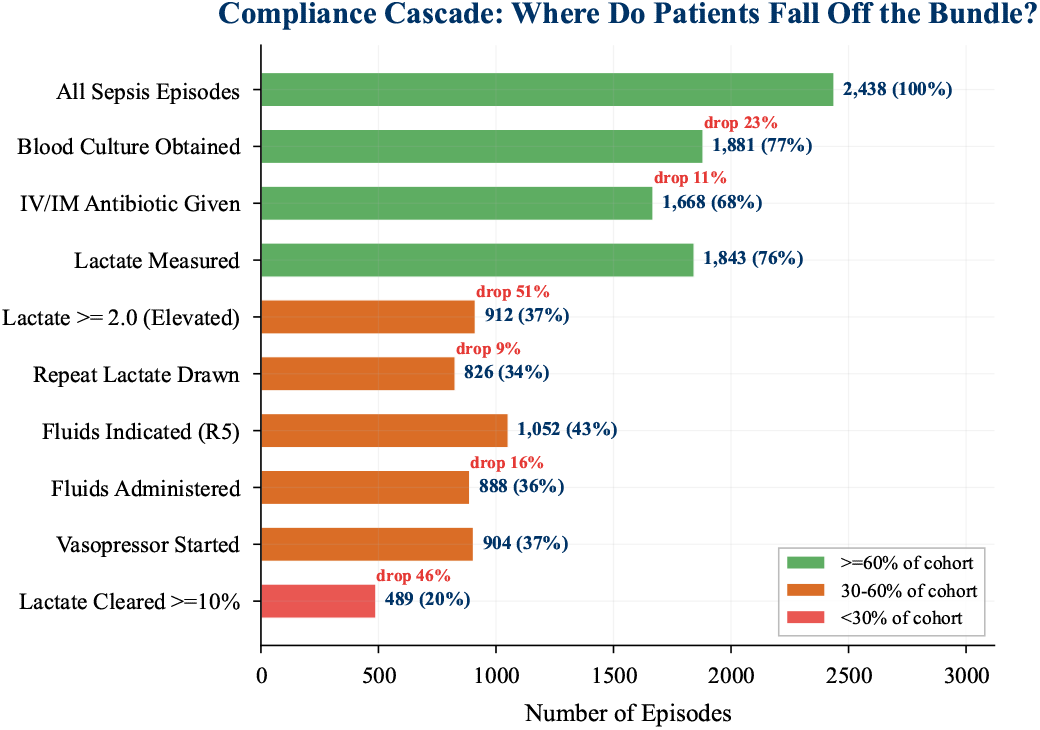
Sequential compliance cascade showing patient attrition across all ten SSC bundle steps for 2,438 sepsis episodes, color-coded by cohort retention tier (≥ 60% green; 30–60% orange; *<*30% red). The most clinically alarming drop occurs at the elevated lactate threshold (≥ 2.0 mmol/L), where only 912 episodes (37%) recorded an elevated value — a 51% fall from those with any lactate measured — signalling widespread under-measurement or under-documentation of hyperlactatemia, a mandatory trigger for SSC resuscitation escalation. Cumulative attrition reduces measurable lactate clearance (≥ 10%) to only 20% of all episodes.

Hour-1 interventions. Whether these gaps reflect care that was never delivered or care that was delivered but never recorded, the pipeline conservatively treats both scenarios as non-compliance, consistent with the SSC guideline philosophy that undocumented care cannot be verified and therefore cannot be credited. Downstream conditional rules further reflect this data sparsity. Fluid resuscitation (R5) activated in only 43% of episodes, and lactate clearance assessment was possible in only 34% of cases (Figure 13). Vasopressor initiation, despite its high compliance score among triggered episodes, was relevant in only 37% of the cohort. These activation rates underscore that the bundle functions as an increasingly narrow funnel, where each successive step depends on the successful completion and documentation of prior steps. The cumulative effect is that only 20% of all sepsis episodes achieved measurable lactate clearance of 10% or more, the final step in the cascade, representing a dramatic attrition from the initial cohort (This answers RQ2; see Appendix: Research Question Summary). The clinical significance of these compliance patterns becomes most apparent when examining their relationship with ICU outcomes (Figure 8). Episodes in the high-compliance tertile achieved a median ICU length of stay of 3.8 days compared to 5.1 days for episodes in the low-compliance tertile, a difference of 1.3 days that carries substantial implications for both patient welfare and health-care resource utilization. The broader trend across all 2,362 episodes with available length of stay data shows a negative correlation (*r* = − 0.026) between compliance score and ICU duration, with an estimated reduction of 0.006 days for each percentage point gain in compliance (Figure 16). While this correlation is modest, the antibiotic timing analysis provides more compelling evidence of clinical impact. Episodes where antibiotics were administered within 30 to 60 minutes of sepsis onset achieved a median ICU stay of just 2.95 days, whereas delays beyond six hours were associated with stays of 4.74 days, representing a 61% increase in median ICU duration. This dose-response relationship between antibiotic delay and ICU length of stay provides strong observational support for the SSC emphasis on rapid antimicrobial therapy as a cornerstone of sepsis management (This answers RQ3; see Appendix: Research Question Summary).

**Fig. 8:**
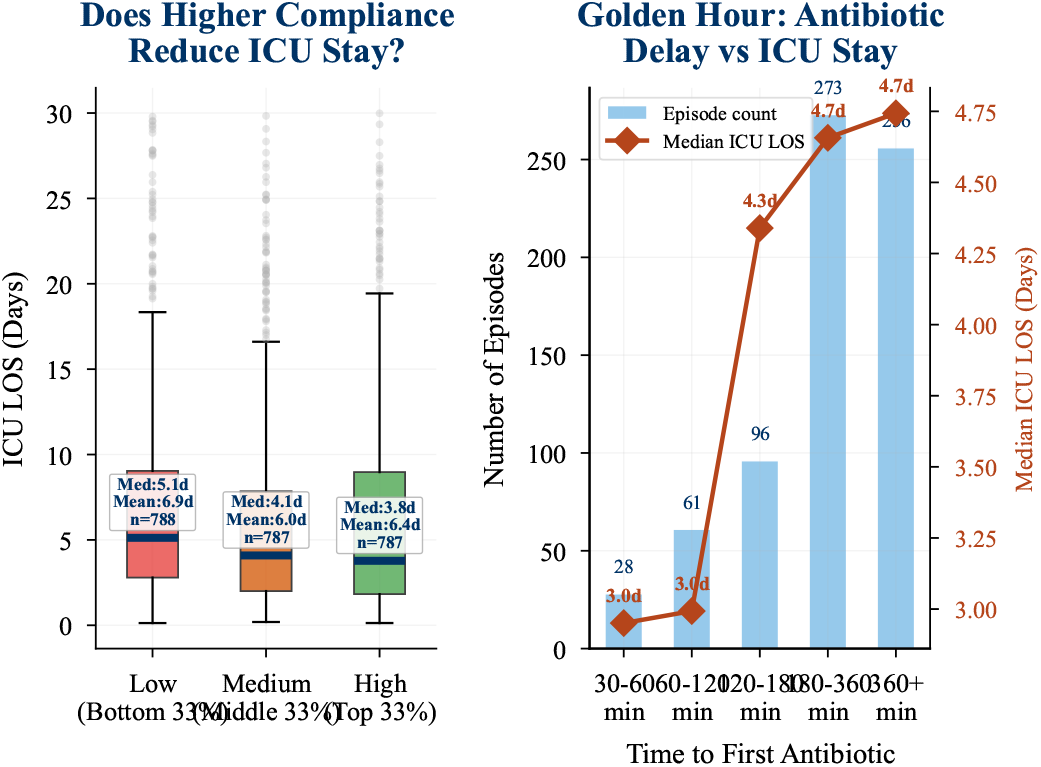
**Left:** ICU length of stay by compliance tertile; high-compliance episodes achieved a median of 3.8 days versus 5.1 days for low-compliance episodes, a 1.3-day difference with direct implications for critical care resource utilization. **Right:** Antibiotic timing dose-response showing median ICU stay rising monotonically from 2.95 days (30–60 min) to 4.74 days (*>*6 hours), a 61% increase providing observational evidence supporting the SSC mandate for rapid empiric antimicrobial therapy in sepsis.

Further analysis of worst-performing rules per episode (Figure 17) localizes the clinical burden to specific bundle components. Episodes where antibiotic timing was the weakest link carried a median ICU stay of 4.7 days, while those bottlenecked by lactate measurement failures had a median of 5.0 days. These two Hour-1 bundle elements together account for the overwhelming majority of episodes and represent the primary drivers of prolonged critical care. The phase-level compliance analysis (Figure 15) reinforces this pattern, showing that median compliance rises from 0.00 in the Hour-1 bundle to 0.67 in the resuscitation phase and 1.00 in the treatment response phase, confirming that immediate interventions constitute the dominant failure mode (This answers RQ1; see Appendix: Research Question Summary). One additional finding merits clinical attention. Among the 2,000 patients in the study cohort, 14% experienced recurrent sepsis episodes, yet median compliance improved only marginally from 0.362 on the first episode to 0.375 on the second, an increase of just 1.3% (Figure 18). This negligible improvement suggests that neither individual clinical learning nor institutional feedback loops produce meaningful compliance gains across repeated admissions for the same patient. From a quality improvement perspective, this finding implies that simply having prior experience managing a septic patient is insufficient to change adherence behavior, and that systemic interventions targeting documentation practices, clinical workflows, and real-time decision support may be necessary to close the compliance gap identified by this pipeline. Taken together, these results paint a consistent picture of sepsis bundle adherence in which the earliest and most time-critical interventions represent the greatest area of failure. The pipeline’s graded fuzzy compliance scores provide more nuanced insight than traditional binary assessments, revealing not just whether a bundle element was completed but how closely its timing and execution aligned with guideline targets. The strong association between higher compliance and shorter ICU stays reinforces the clinical imperative behind the SSC bundle and suggests that even incremental improvements in Hour-1 adherence could translate into meaningful reductions in critical care resource consumption and patient morbidity.

## VI. CONCLUSION

In this paper, we presented an Expert-Guided Neuro-Symbolic Pipeline that combines LLM-based semantic normalization, domain expert consultation, and a Sugeno fuzzy inference system to produce interpretable, graded compliance assessments for sepsis care across 2,438 episodes from *MIMIC* − *IV* v3.1. Unlike prior work relying on binary compliance judgments at the bundle level, aggregated program-level metrics, or admission-time stratification, our pipeline operates at per-rule, per-episode, and per-phase granularity (see Appendix: Comparison Analysis). The semantic normalization layer resolved 220 clinically relevant drug strings that purely symbolic systems would have missed, achieving 94.26% inter-classifier agreement (*κ* = 0.65) and 98.59% embedding confirmation, while the fuzzy inference system produced graded scores aligned with SSC safety boundaries that no purely neural model could guarantee. The Hour-1 bundle emerges as the dominant clinical failure mode, with antibiotic timing (R2, *μ* = 0.24), blood culture sequencing (R1, *μ* = 0.36), and lactate measurement (R3, *μ* = 0.39) driving mean overall compliance of just 36.7%. The 51% patient drop-off at the elevated lactate threshold (Figure 7) represents a particularly actionable finding pointing to systematic under-measurement or under-documentation amenable to automated order sets and EHR decision support alerts. The association between higher compliance and shorter ICU stays, with 3.8-day median stays for high-compliance versus 5.1 days for low-compliance episodes, and antibiotic timing showing stays rising from 2.95 days at 30-60 minutes to 4.74 days beyond six hours (Figure 8), provides strong observational support for the SSC emphasis on rapid intervention. High conditional-rule performance (R6, *μ* = 0.73; R7, *μ* = 0.97) likely reflects survivorship bias rather than genuine excellence, cautioning against interpreting downstream metrics in isolation. The negligible 1.3% compliance improvement between first and second sepsis episodes (Figure 18) suggests that institutional-level interventions, not individual clinical experience, are necessary to close the adherence gap. Several limitations warrant acknowledgment. First, the pipeline depends on domain experts for decision boundaries and rule parameterization, potentially limiting scalability in resource-constrained settings. Second, the analysis is retrospective and single-center (*MIMIC* −*IV*, Beth Israel Deaconess Medical Center), and compliance patterns may not generalize across institutions with different documentation practices and EHR systems. Third, timing measurements anchored to the recorded sepsis onset timestamp penalize pre-ICU antibiotic administration that may represent appropriate early action (Figure 12). Fourth, treating missing data as non-compliance, while consistent with SSC philosophy, cannot distinguish undelivered care from undocumented care. Fifth, membership function parameters require fresh expert input for each new clinical protocol. This pipeline is not limited to sepsis and can be adapted to any protocol with well-defined time-sensitive interventions, such as stroke management, acute myocardial infarction care, or trauma resuscitation. Looking forward, as generative AI models mature in medical reasoning, they may progressively reduce dependency on manual expert consultation, enabling more autonomous clinical decision support. Multi-center validation, real-time EHR integration, and prospective evaluation of whether graded compliance feedback improves clinical outcomes (seeAppendix: Quality of Life Implications) represent the most important next steps for translating this work into actionable clinical tools.

## Data Availability

The datasets analyzed during the current study are available from the MIMIC-IV v3.1 repository on PhysioNet. Access is controlled and requires completion of PhysioNet credentialing, required training, and acceptance of the data use agreement. The database is de-identified and available only to approved users.

https://physionet.org/content/mimiciv/3.1/

## APPENDIX

### A. Data Availability and Ethical Statement

This study utilizes the MIMIC-IV v3.1 database, a deidentified critical care dataset derived from the electronic health records of Beth Israel Deaconess Medical Center (BIDMC). The database covers the period from 2008 to 2022 and includes information on roughly 250,000 patients across about 500,000 hospital admissions. MIMIC-IV is publicly available through PhysioNet [5]; however, it is distributed under a credentialed-access model. Researchers must complete approved human subjects research training and sign a PhysioNet Credentialed Health Data Use Agreement (DUA), which restricts attempts to re-identify individuals, prohibits redistribution of the dataset, and limits its use strictly to research purposes. The BIDMC Institutional Review Board (IRB) approved the release of MIMIC-IV as a research resource and granted a waiver of informed consent. As a result, no additional IRB approval was required for this study. All personally identifiable information was removed prior to release following the HIPAA Safe Harbor de-identification standards. Due to the data use agreement, the raw MIMIC-IV dataset, derived cohorts, and episode-level files generated for this study cannot be distributed by the authors and must instead be obtained directly from PhysioNet.

### B. Eight Rules From SSC

The compliance evaluation within this pipeline is dictated by eight specific guidelines derived from the Surviving Sepsis Campaign (SSC) [1], outlined below:

1. [*R*1**] Blood Cultures Prior to Antimicrobials:**Secure samples for pathogen identification before the blood-stream is sterilized by treatments. *“Obtain blood cultures before administering antibiotics*.*”*
2. [*R*2**] Empiric Broad-Spectrum Antibiotics:**Promptly commence broad-spectrum antimicrobial therapy to ensure immediate coverage. *“Administer broad-spectrum antibiotics*.*”*
3. [*R*3**] Initial Lactate Measurement:**Evaluate the extent of tissue hypoperfusion as soon as sepsis is recognized. *“Measure lactate level*.*”*
4. [*R*4**] Lactate Re-evaluation:**Verify ongoing hypoperfusion if the baseline lactate measurement is abnormally high. *“Remeasure lactate if initial lactate is elevated (> 2 mmol/L)*.*”*
5. [*R*5**] Intravenous Fluid Therapy:**Replenish intravascular volume for patients experiencing hemodynamic instability. *“Begin rapid administration of 30 mL/kg crystalloid for hypotension or lactate* ≥ *4 mmol/L*.*”*
6. [*R*6**] Vasopressor Administration:**Augment perfusion pressure when intravenous fluids alone fail to stabilize the patient. *“Apply vasopressors if hypotensive during or after fluid resuscitation to maintain a mean arterial pressure (MAP)* ≥ *65 mm Hg*.*”*
7. [*R*7**] Mean Arterial Pressure (MAP) Goal:**Aim for a baseline perfusion threshold to mitigate end-organ ischemia. *“For adults with septic shock on vasopressors, we recommend an initial target mean arterial pressure (MAP) of 65 mm Hg over higher MAP targets*.*”*
8. [*R*8**] Monitoring Lactate Clearance:**Confirm a positive metabolic trajectory in response to ongoing resuscitation therapies. *“For adults with sepsis or septic shock, we suggest guiding resuscitation to decrease serum lactate in patients with elevated lactate levels over not using serum lactate*.*”*

**Note:** The 10% threshold for lactate clearance referenced in Rule 8 is not a direct component of the SSC guidelines; rather, it is adapted from the research of Jones et al. [28].

### C. Research Question Summary

- **[RQ1] Which specific SSC bundle interventions represent the most critical points of clinical failure in sepsis management, and how does a graded compliance framework reveal failure patterns that binary assessment cannot capture?** The Sugeno fuzzy inference system reveals stark stratification across eight bundle rules, with antibiotic timing (R2, *µ* = 0.24) as the most critical compliance failure, followed by blood culture sequencing (R1, *µ* = 0.36) and lactate measurement (R3, *µ* = 0.39), all falling below the 50% threshold. Conditional hemodynamic rules such as vasopressor initiation (R6, *µ* = 0.73) and MAP recovery (R7, *µ* = 0.97) appear near-acceptable but reflect survivorship bias rather than genuine clinical excellence. The graded scoring enabled phase-level analysis showing median compliance rising from 0.00 in the Hour-1 bundle to 0.67 in resuscitation and 1.00 in treatment response, confirming that immediate interventions constitute the dominant failure mode and exposing patterns that binary compliance systems could not surface.
- **[RQ2] Where do patients fall off the sequential sepsis management pathway, and what do the cascade drop-off patterns reveal about systemic gaps in early sepsis intervention and clinical documentation?** The compliance cascade reveals that 77% of episodes had blood cultures obtained and 76% had lactate measured, but a striking 51% patient drop-off occurs at the elevated lactate threshold (≥ 2.0 mmol/L), with only 912 episodes (37%) recording this value. Antibiotic timing (31.6% missing) and lactate measurement (24.4% missing) are the most data-sparse Hour-1 interventions. Downstream, fluid resuscitation activated in only 43% of episodes and lactate clearance assessment was possible in only 34% of cases, with the cumulative effect that only 20% of all episodes achieved measurable lactate clearance of 10% or more. The pipeline conservatively treats both undelivered and undocumented care as non-compliance, consistent with the SSC guideline philosophy that unverifiable care cannot be credited.
- **[RQ3] How does temporal adherence to SSC bundle components, particularly antibiotic timing and overall compliance, correlate with ICU length of stay and patient outcomes?** Episodes in the high-compliance tertile achieved a median ICU length of stay of 3.8 days compared to 5.1 days for the low-compliance tertile, with a broader trend showing a reduction of 0.006 days per 1% compliance gain (*r* = −0.026, *n* = 2,362). The antibiotic timing dose-response relationship shows median ICU stays rising from 2.95 days at 30 to 60 minutes to 4.74 days beyond six hours, a 61% increase. Worst-rule analysis localizes the highest ICU burden to antibiotic timing (R2, median 4.7 days) and lactate measurement (R3, median 5.0 days), establishing Hour-1 intervention failures as the primary driver of prolonged critical care. Additionally, recurrent sepsis patients showed only a 1.3% compliance improvement between episodes, indicating that repeated clinical exposure alone does not improve adherence.

### D. Data Overview

The data pipeline processes information from the *MIMIC* − *IV v*3.1 [5] database, an extensive critical care electronic health record repository covering the years 2008 to 2022. It encompasses roughly 250, 000 unique patients and over 500, 000 distinct hospital admissions. The database is partitioned into two core relational modules: *MIMIC* −*IV/hosp* for hospital-wide clinical and administrative records, and *MIMIC*− *IV/icu* for granular intensive care unit events and measurements. As illustrated in Figure 9, data integration across different clinical granularities is achieved using three primary keys:

**Fig. 9:**
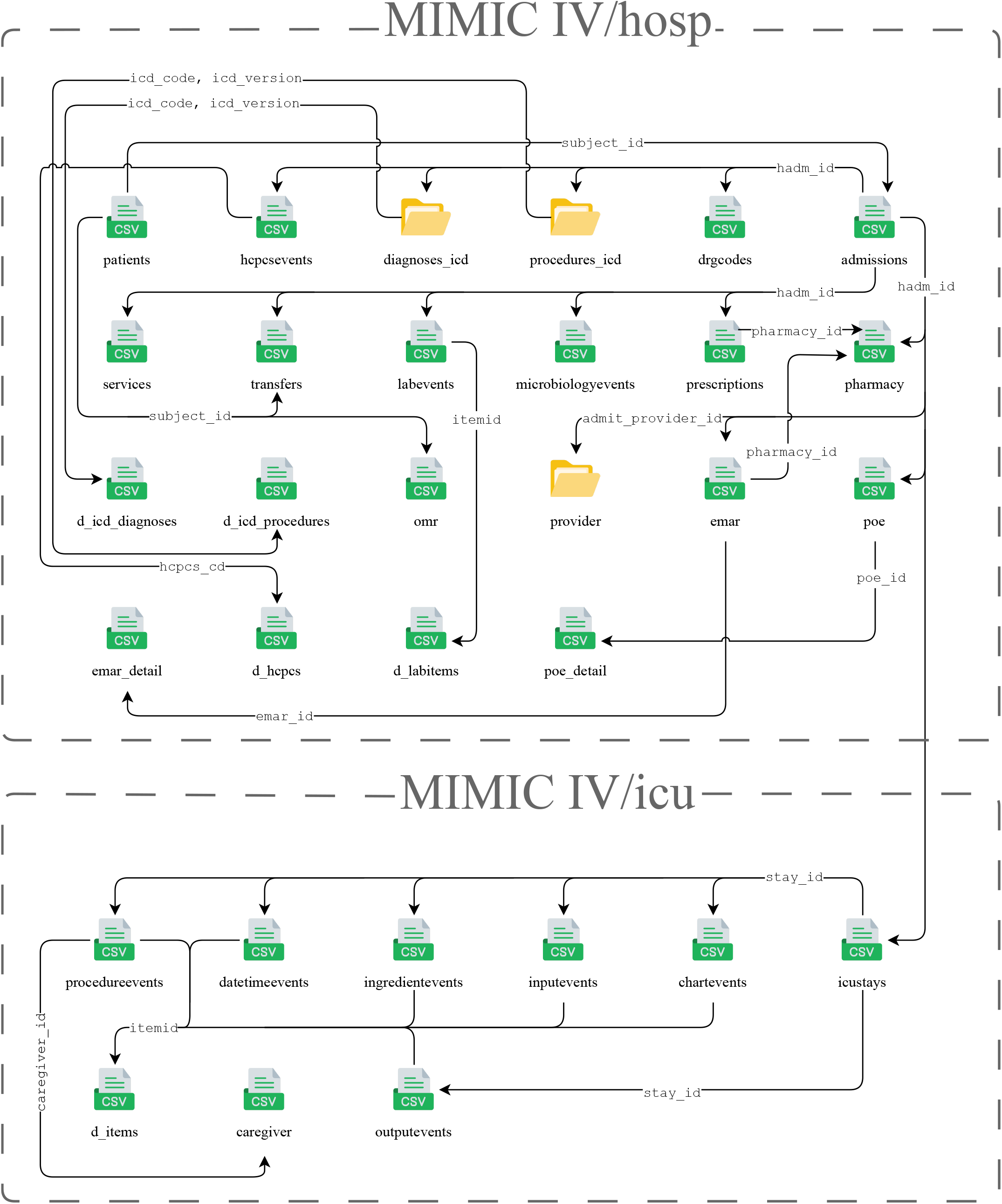
MIMIC-IV Database Structure and Relational Organization. The database comprises two modules: MIMIC-IV/hosp maintaining hospital-level records indexed by `subject_id`, `hadm_id`, and `pharmacy_id`, and MIMIC-IV/icu (9 tables, bottom portion) maintaining ICU-specific records indexed by `stay_id` . Primary linking identifiers (`subject_id`, `hadm_id`, `stay_id`) enable integration across administrative, clinical, and intensive care data layers. The pipeline extracts data from `emar`, `prescriptions`, `microbiologyevents`, `labevents`, `inputevents`, `chartevents`, and `icustays` .

**Fig. 10:**
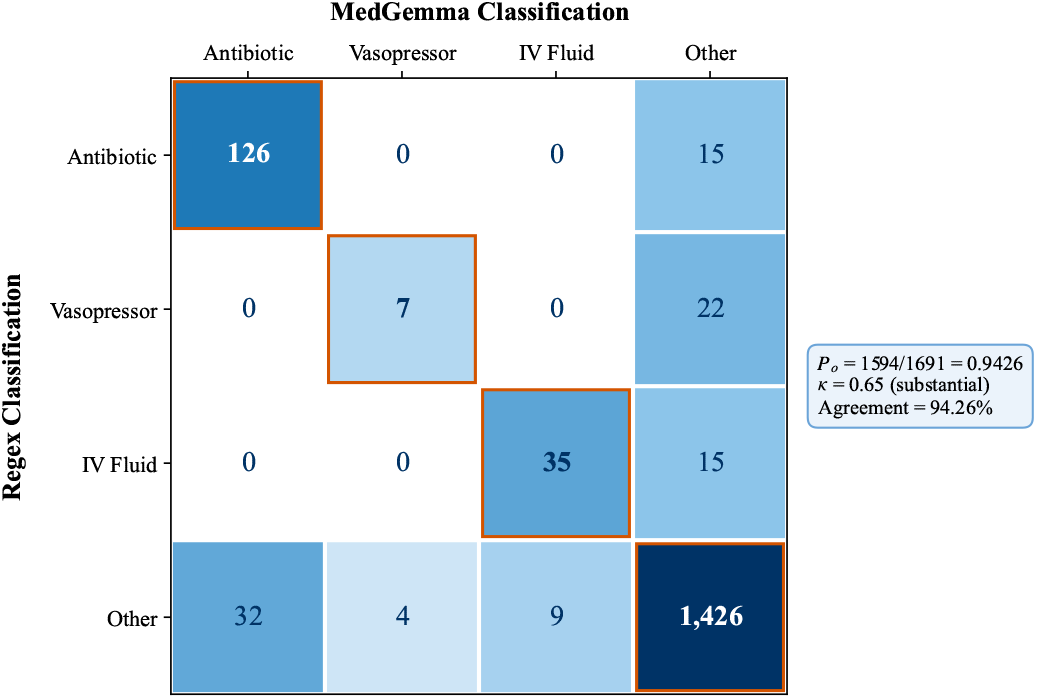
Drug classification confusion matrix comparing regex-based and MedGemma-4b-it classifiers across all 1,691 unique drug strings extracted from MIMIC-IV electronic health records. Diagonal cells (orange borders) represent inter-classifier agreement (*P*_*o*_=1,594*/*1,691=0.9426; *κ*=0.65, substantial). All off-diagonal disagreements within the clinically critical submatrix (antibiotic, vasopressor, IV fluid) are zero, confirming that when classifiers disagree, the error is always between a clinical category and *other* — never a cross-clinical misclassification (e.g., a vasopressor mistaken for an antibiotic) that could catastrophically distort bundle compliance scores. Regex misses 32 antibiotics, 4 vasopressors, and 9 IV fluids that MedGemma captures (synonyms and trade names); conversely, MedGemma misses 15 antibiotics, 22 vasopressors, and 15 IV fluids that regex captures. This complementary error profile, confirmed by McNemar’s test (*p*=0.088), justifies the dual-classifier consensus architecture for clinically safe drug normalization.

**Fig. 11:**
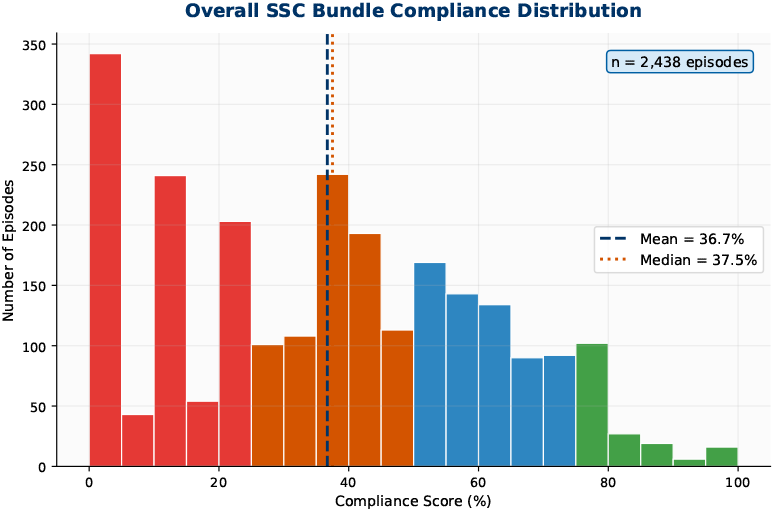
Frequency distribution of overall SSC bundle compliance scores across all 2,438 sepsis episodes, color-coded by compliance quartile. The distribution is markedly left-skewed, with a mean of 36.7% and median of 37.5%, indicating that the majority of episodes fall in the lowest performance tiers despite representing real-world intensive care practice at a major academic medical center. The heavy concentration of episodes below 50% compliance reflects systemic failure in Hour-1 bundle execution, particularly antibiotic timing, blood culture sequencing, and initial lactate measurement, rather than random variation in care quality, and highlights the population-level burden of suboptimal early sepsis management.

**Fig. 12:**
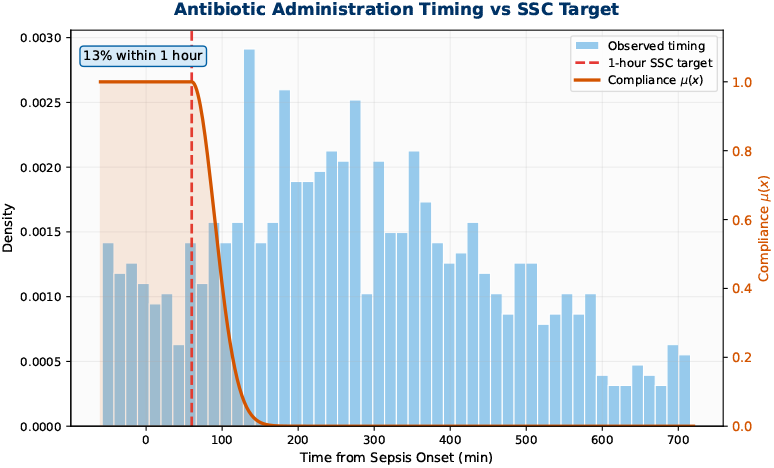
Distribution of time elapsed from sepsis onset to first IV/IM antibiotic administration (teal histogram) across episodes with available timing data, overlaid with the fuzzy compliance membership function *µ*(*x*) (orange curve, right y-axis). The dashed red vertical line marks the 1-hour SSC target. Only 13% of episodes met this strict mandate, with the distribution peaking at approximately 130–180 minutes post-onset. The membership function decay (half-Gaussian, *c*=60 min, *σ*=30) captures the clinically established principle that antimicrobial benefit degrades continuously with delay rather than abruptly at the 60-minute boundary. The leftward tail of the histogram, where administration time is negative (antibiotics given before the recorded sepsis onset), reflects pre-ICU antibiotic initiation, a common real-world workflow where empiric treatment begins in the emergency department prior to formal sepsis recognition and represents a structural limitation of retrospective timestamp-anchored compliance analysis.

**Fig. 13:**
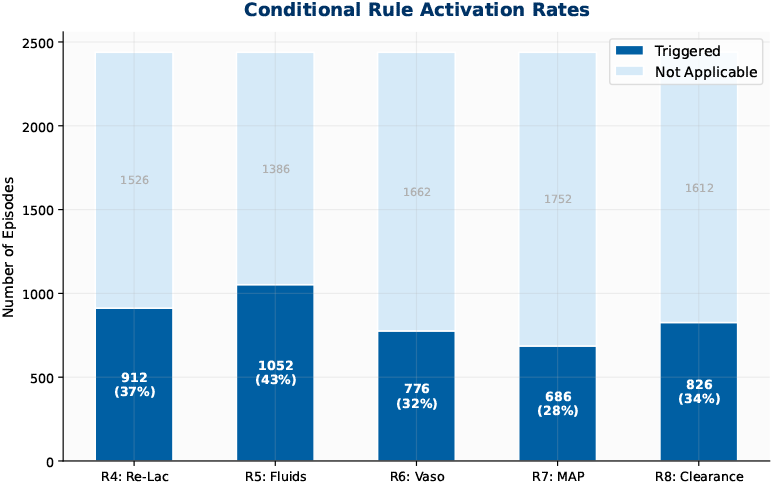
Stacked bar chart displaying the proportion of the 2,438-episode cohort triggering each of the five conditional SSC bundle rules (R4–R8), reflecting the clinical acuity distribution within the cohort. Fluid resuscitation (R5) was the most frequently activated conditional intervention (1,052 episodes; 43%), indicating a high prevalence of hemodynamic instability (MAP *<*65 mmHg) or severe hyperlactatemia (lactate *≥* 4 mmol/L). Repeat lactate assessment (R4) was triggered in 912 episodes (37%), all of whom had documented initial lactate *≥* 2.0 mmol/L. Vasopressor initiation (R6) applied to 776 episodes (32%), MAP recovery assessment (R7) to 686 episodes (28%), and lactate clearance evaluation (R8) to 826 episodes (34%). These activation rates underscore that the SSC bundle functions as a progressively narrowing clinical funnel, where each downstream rule is evaluated in an increasingly acutely ill subset of the original cohort.

**Fig. 14:**
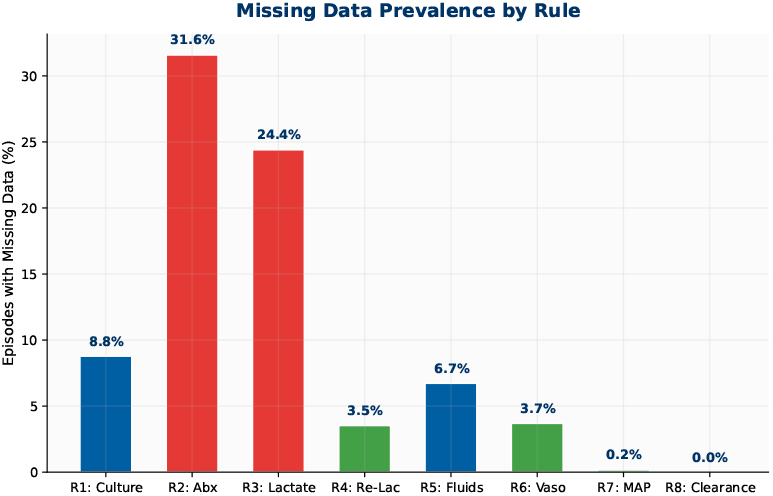
Prevalence of missing clinical documentation across the eight SSC bundle components, expressed as the percentage of applicable episodes lacking the data required for rule evaluation. Data sparsity is most severe for the two most time-sensitive Hour-1 interventions: antibiotic administration timing (R2, 31.6% missing) and initial lactate measurement timing (R3, 24.4% missing). Blood culture documentation (R1, 8.8% missing) and fluid administration records (R5, 6.7% missing) exhibit moderate sparsity. Vasopressor-related rules (R6, 3.7%; R4, 3.5%) and hemodynamic response metrics (R7, 0.2%; R8, 0.0%) are near-complete. Consistent with SSC guideline philosophy, that unverifiable care cannot be credited, the pipeline conservatively scores all missing mandatory rule data (R1-R3) as non-compliant (*µ*=0), acknowledging that it is impossible to distinguish undelivered care from undocumented care in retrospective EHR analysis.

**Fig. 15:**
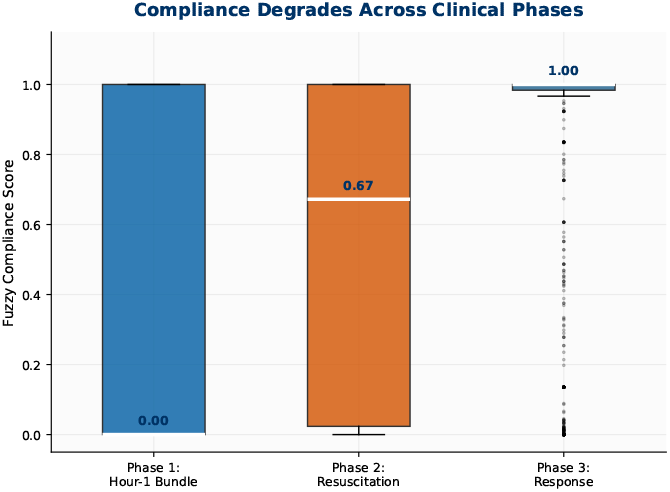
Box plots illustrating the distribution of fuzzy compliance scores stratified by clinical care phase: Phase 1 (Hour-1 Bundle: R1-R3), Phase 2 (Hemodynamic Resuscitation: R5-R6), and Phase 3 (Treatment Response: R7-R8). Median compliance rises markedly across phases: 0.00 in the Hour-1 bundle, 0.67 in resuscitation, and 1.00 in treatment response. The near-zero median for Phase 1 reflects the dominant failure mode identified in this study, simultaneous underperformance in blood culture sequencing, antibiotic timing, and lactate measurement, which collectively drive mean overall bundle compliance to just 36.7%. The apparent improvement in downstream phases must be interpreted cautiously: Phase 2 and Phase 3 rules activate only in patients who survived to require secondary intervention, introducing survivorship bias that inflates conditional compliance scores relative to the full septic cohort. The trend line looks flat because of the possible outliers, which we did have information of what to remove.

**Fig. 16:**
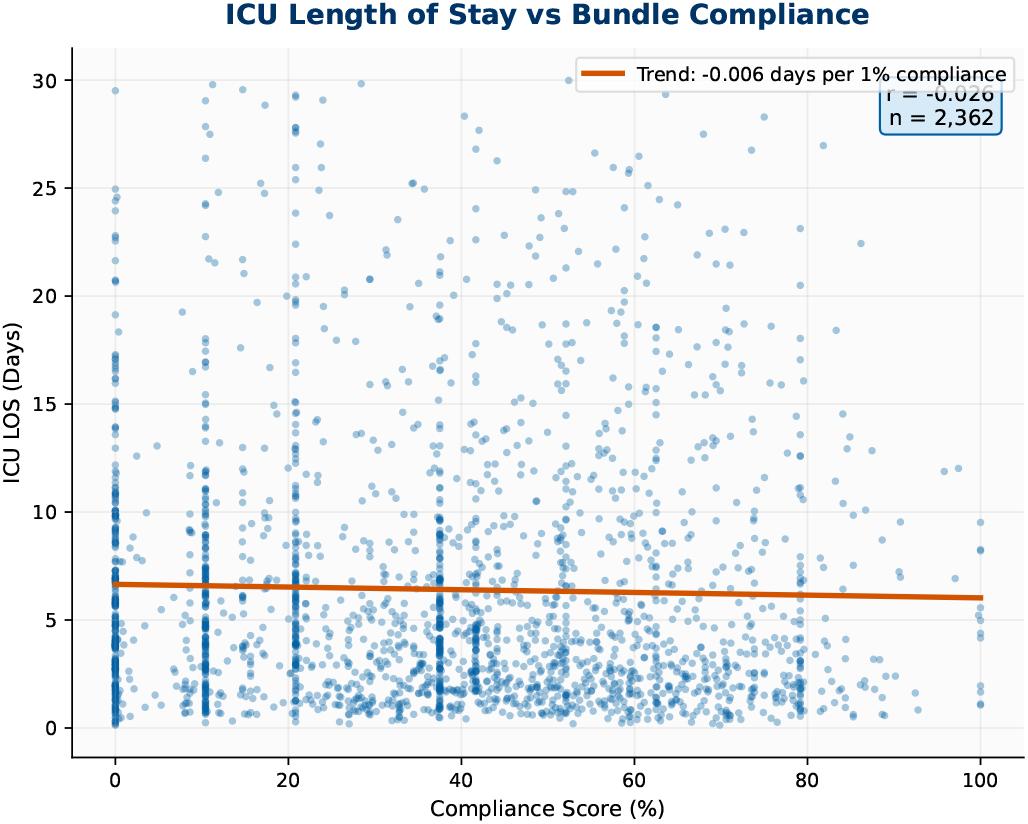
Scatter plot evaluating the relationship between overall SSC bundle compliance score and ICU length of stay (LOS) across 2,362 episodes with complete outcome data. Each point represents one sepsis episode. The orange regression line demonstrates a negative trend of*−* 0.006 days of ICU stay per 1% increase in compliance (*r*= *−*0.026, *n*=2,362). Although the correlation is modest, reflecting substantial clinical heterogeneity in sepsis severity, comorbidity burden, and ICU admission indication, the directional relationship is consistent with the causal model underlying SSC guidelines. The weak aggregate correlation contrasts with the stronger dose-response signal observed for antibiotic timing specifically (Figure 12 / Figure 8), suggesting that the compliance-outcome relationship is concentrated in the Hour-1 bundle rather than distributed uniformly across all eight rules. Readers should note that this correlation reflects a single academic center, Beth Israel Deaconess Medical Center (BIDMC) and is subject to timestamp anchoring bias, whereby pre-ICU antibiotic administration is scored as non-compliant, and to survivorship bias in the triggered subset. Both of which likely suppress the true compliance outcome gradient and caution against direct generalization to institutions with different documentation practices or patient populations.

**Fig. 17:**
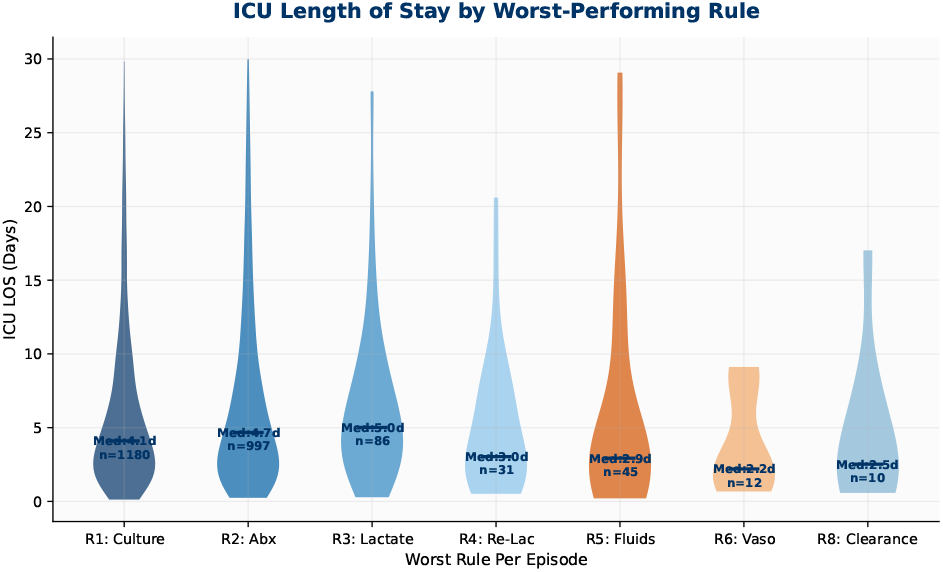
Violin plots displaying ICU length of stay distributions grouped by the SSC bundle rule that yielded the lowest per-episode fuzzy compliance score (the “worst-performing” rule), providing a clinician-actionable view of which bundle failures impose the greatest burden on critical care resources. Episodes where antibiotic timing (R2) was the weakest link carried a median ICU stay of 4.7 days (*n*=997), and those bottlenecked by initial lactate measurement failure (R3) had a median of 5.0 days (*n*=86). Together, these two Hour-1 bundle components account for the overwhelming majority of episodes and represent the primary drivers of prolonged ICU admission. By contrast, episodes limited by fluid resuscitation (R5, median 2.9 days) or vasopressor compliance (R6, median 2.2 days) tended toward shorter stays, consistent with the interpretation that patients who successfully complete early interventions and are bottlenecked only at downstream rules have experienced more clinically effective initial management.

**Fig. 18:**
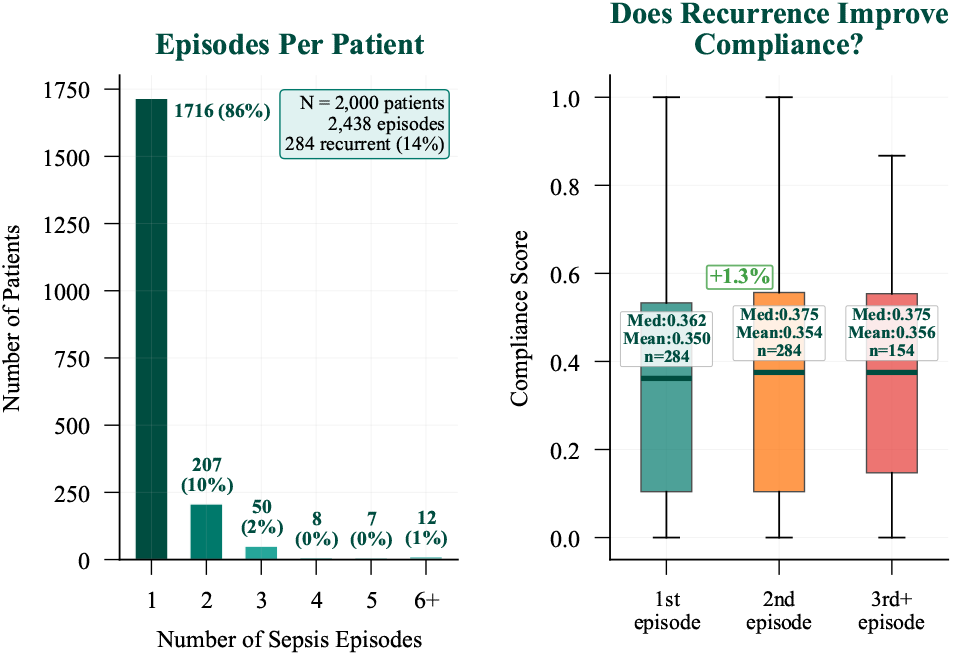
**Left - Recurrence Distribution:** Bar chart showing the distribution of sepsis episode frequency per patient across the 2,000-patient cohort (2,438 total episodes). The majority of patients (1,716; 86%) experienced a single sepsis hospitalization, while 284 patients (14%) had recurrent episodes, with a small subset experiencing three or more admissions, a pattern associated with immunosuppression, chronic comorbidities, and indwelling devices. **Right - Recurrence and Compliance:** Box plots comparing SSC bundle compliance scores between first (*n*=284; median 0.362), second (*n*=284; median 0.375), and third-or-later (*n*=154; median 0.375) episodes in recurrent patients. The marginal 1.3% improvement in median compliance between first and second encounters indicates that individual clinical exposure to sepsis management does not translate into meaningful adherence gains, suggesting that systemic institutional interventions, including automated EHR order sets, real-time decision support alerts, and structured documentation workflows, rather than experiential learning alone, are required to close the sepsis bundle compliance gap.

1. `subject_id` (identifies the unique patient)
2. `hadm_id` (identifies the unique hospital admission)
3. `stay_id` (identifies the unique ICU stay)

The *MIMIC* − *IV/hosp* module is hierarchically structured into 22 tables. General demographic and admission data are stored in the `patients` and `admissions` tables. Clinical records linked by `hadm_id` capture coded medical data, laboratory testing, and pharmacology. Standardized conceptual mappings for these records are provided by reference tables (`d_icd_diagnoses`, `d_icd_procedures`, `d_hcpcs`, `d_labitems`). The clinical tables include:

1. `diagnoses_icd` and `procedures_icd` (coded diagnoses and procedures)
2. `labevents` (laboratory measurements)
3. `microbiologyevents` (culture results)
4. `prescriptions` and `pharmacy` (medication records)
5. `emar` and `poe` (medication administration events at granular timestamps)

Conversely, the *MIMIC* −*IV/icu* module consists of 9 tables linked by `stay_id` . ICU admissions are grounded by the `icustays` table. The `d_items` dictionary table translates `itemid` codes into human-readable clinical concepts. Measurement and event data are distributed across the following tables:

1. `chartevents` (bedside charted observations)
2. `inputevents` (administered medications and fluids)
3. `outputevents` (fluid excretion)
4. `procedureevents` (performed procedures)
5. `datetimeevents` (timestamped clinical milestones)
6. `ingredientevents` (granular medication components)

To evaluate compliance with the Surviving Sepsis Campaign (SSC), the extraction pipeline targets seven essential tables across both modules to fulfill six specific bundle requirements:

1. **Antibiotics:** Sourced from medications.
2. **Vasopressors:** Sourced from medications.
3. **IV fluids:** Tracked via `inputevents` .
4. **Blood cultures:** Procured from `microbiologyevents` .
5. **Lactate measurement:** Retrieved from `labevents` .
6. **Hemodynamic targets:** Monitored through vital signs in `chartevents` .

These clinical indicators are specifically derived from the following sources:

1. **From** *MIMIC* −*IV/hosp*: `prescriptions` and `emar` provide medication names and administration schedules; `microbiologyevents` supplies interpreted organisms and specimen types; and `labevents` yields lactate levels and other diagnostic assays.
2. **From** *MIMIC* − *IV/icu*: `inputevents` tracks vasopressor and fluid administration; `chartevents` captures blood pressure and associated vital signs; and `icustays` establishes the chronological baseline of the ICU admission.

While this relational architecture successfully facilitates the linkage of patient demographics with longitudinal clinical events across multiple hospitalizations via `subject_id`, `hadm_id`, and `stay_id`, it also introduces challenges regarding data consistency. Specifically, unstructured text is prevalent throughout multiple locations; for instance, medication terminology in the `prescriptions` table inconsistently utilizes abbreviations, generic nomenclature, and trade names. This textual heterogeneity mandates the implementation of the semantic normalization step detailed in Section IV-B.

### E Fuzzy Membership Function Parameter

**TABLE II:**
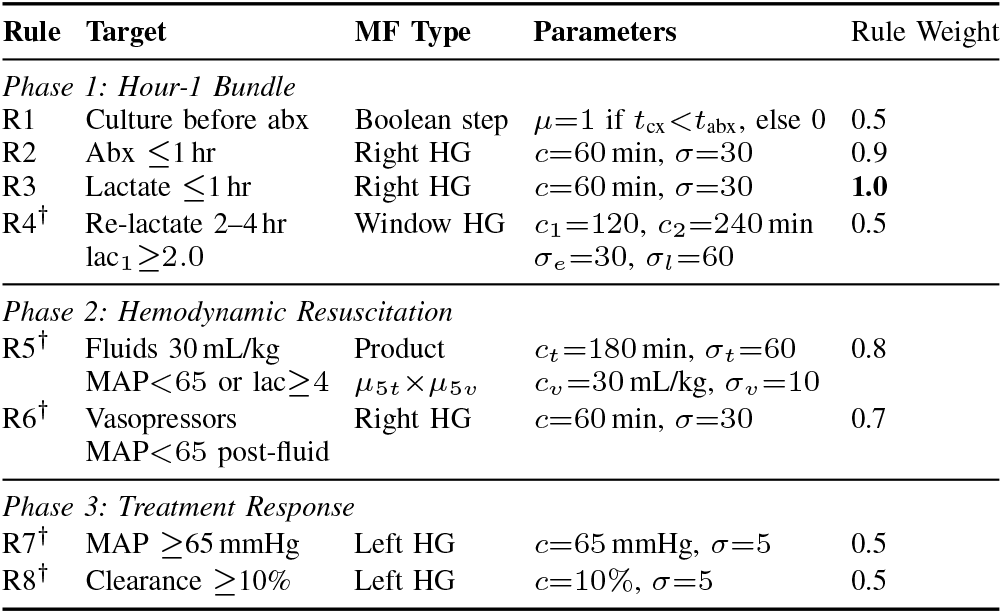
Membership function parameters for all eight SSC bundle compliance rules, with clinical decision boundaries (*c*) and Gaussian decay rates (*σ*) established through domain expert consultation with practicing intensivists and infectious disease specialists. Phase 1 (Hour-1 Bundle) rules encode the most time-sensitive interventions: R1 uses a Boolean step function (blood culture before antibiotics); R2 and R3 apply right-sided half-Gaussians with *c*=60 min and *σ*=30, reflecting the SSC 1-hour mandate and the clinical evidence that antimicrobial and lactate measurement benefit degrades gradually beyond this threshold; R4 uses a window function (*c*_1_=120 min, *c*_2_=240 min) penalizing both premature (*σ*_*e*_=30) and delayed (*σ*_*l*_=60) repeat lactate sampling. Phase 2 (Resuscitation) rules encode hemodynamic rescue thresholds (R5: 30 mL/kg fluid target; R6: vasopressor within 60 minutes of fluid failure). Phase 3 (Response) rules encode physiologic recovery targets (R7: MAP≥65 mmHg; R8: lactate clearance≥10%). Conditional rules (†) activate only when specified hemodynamic or metabolic triggers are documented. Rule weights encode the SME-established clinical priority ordering, with lactate measurement (R3, weight 1.0) and antibiotic timing (R2, weight 0.9) weighted most heavily.

**TABLE III:** Sugeno fuzzy inference system properties and missing data handling protocol, implementing the SSC guideline philosophy that undocumented care cannot be credited toward compliance. The Sugeno first-order inference type enables computationally efficient weighted aggregation of per-rule outputs via the defuzzification formula 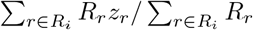, where *R*_*r*_ encodes expert-validated rule weights and *z*_*r*_ the per-rule fuzzy output. Conditional rules that are clinically not applicable (trigger condition unmet) receive *µ*=1.0 and are excluded from the episode’s active rule set *R*_*i*_, preventing inappropriate penalization of patients who did not require hemodynamic escalation. Mandatory Hour-1 rules (R1-R3) with missing documentation receive *µ*=0, conservatively treating data absence as non-compliance. Special handling is applied for R1 when a blood culture is obtained without a subsequent positive pathogen (scored *µ*=0.5) and for R5 when patient weight is unavailable for mL/kg calculation (*µ*_5*v*_=0.5 penalty). Rule priority ordering (*R*_3_*>R*_2_*>R*_5_*>R*_6_*>R*_1_=*R*_4_=*R*_7_=*R*_8_) reflects clinical consensus on the relative mortality impact of individual SSC interventions.

| Property | Value |
| --- | --- |
| Inference type | Sugeno first-order |
| Defuzzification | $\sum_{r \in R_i} R_r z_r / \sum_{r \in R_i} R_r$ |
| Conditional untriggered | $\mu = 1.0$, excluded from $R_i$ |
| Missing mandatory (R1–R3) | $\mu = 0$ |
| Culture only, no abx (R1) | $\mu = 0.5$ |
| No weight available (R5) | $\mu_{5v} = 0.5$ penalty |
| Missing trigger data | Excluded from $R_i$ |
| Classification | Consensus: agree ∪ (embed $\geq \theta$) ∪ SME |

### F. Drug classification Confusion Matrix

### G. Additional Graphs

### H. Comparison Analysis

**TABLE IV:**
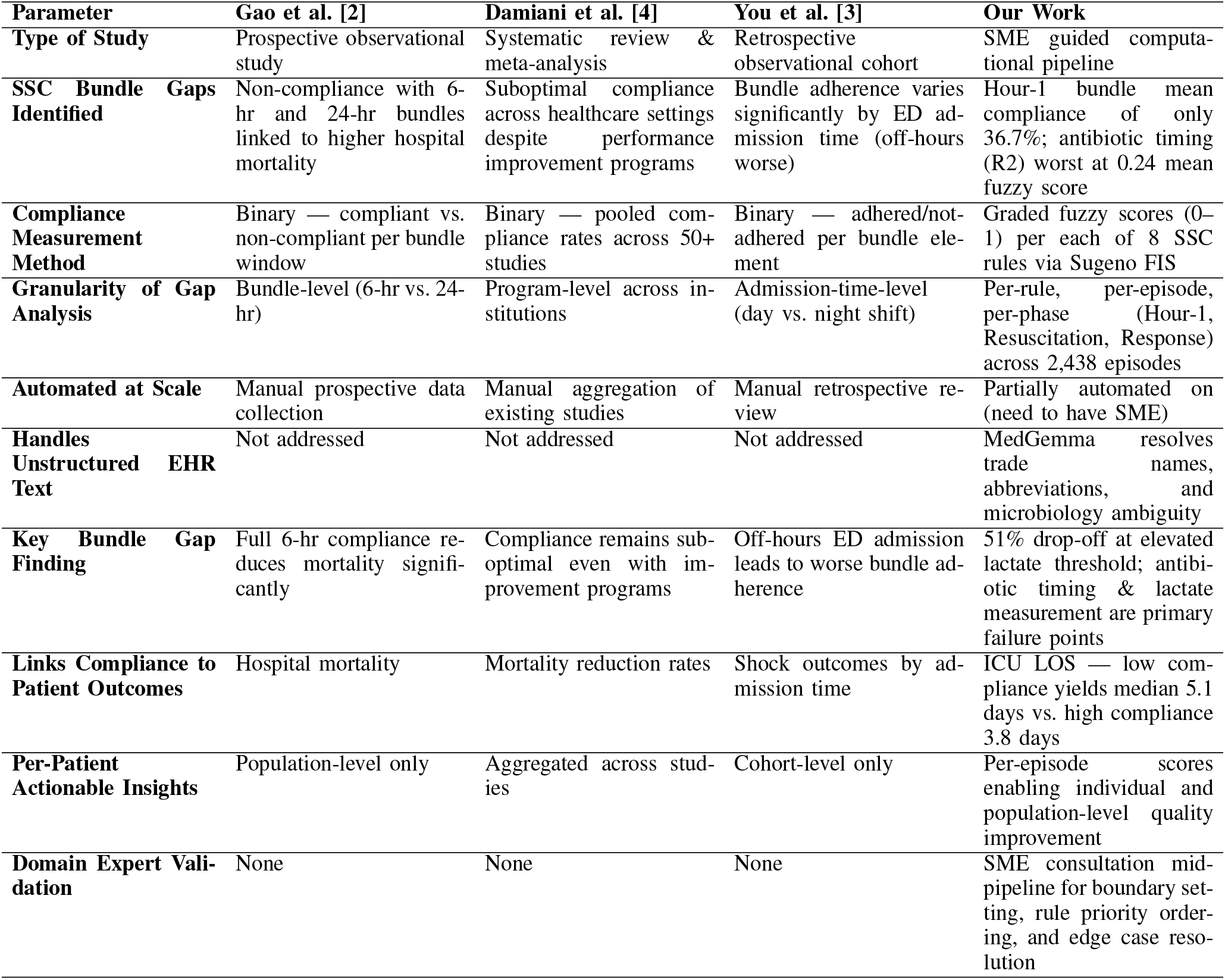
Methodological comparison of the proposed Expert-Guided Neuro-Symbolic Pipeline against three landmark SSC bundle compliance studies. Gao et al. assessed 6-hour and 24-hour bundle compliance prospectively in a single cohort, demonstrating mortality reduction but limited to binary bundle-level judgments. Damiani et al. conducted a systematic review and meta-analysis of compliance rates across institutions, providing population-level evidence of persistent suboptimality but without individual episode granularity or automated EHR processing. You et al. performed a retrospective cohort study revealing that off-hours emergency department admissions are associated with worse bundle adherence, but relied on manual review and binary rule evaluation. The proposed pipeline advances beyond all three antecedents by providing graded per-rule, per-episode, and per-phase compliance scores via a Sugeno fuzzy inference system; automated semantic normalization of unstructured EHR drug names and microbiology results using MedGemma; domain expert consultation integrated mid-pipeline for boundary setting and edge case resolution; and direct linkage of compliance scores to individual patient ICU outcomes at scale across 2,438 episodes, enabling actionable quality improvement at both population and patient levels.

### I. Combined Results

**TABLE V:** Comprehensive summary of quantitative results from the Expert-Guided Neuro-Symbolic Pipeline applied to 2,438 sepsis episodes across 2,000 patients from MIMIC-IV v3.1. Key normalization metrics confirm pipeline reliability: 94.26% inter-classifier drug agreement (*κ*=0.65) and 98.59% embedding-based validation rate. Rule-level compliance scores reveal that antibiotic timing (R2, *µ*=0.24) and blood culture sequencing (R1, *µ*=0.36) are the most critical Hour-1 failures, while vasopressor initiation (R6, *µ*=0.73) and MAP recovery (R7, *µ*=0.97) reflect survivorship bias in the triggered subset. The compliance cascade identifies a 51% patient drop-off at the elevated lactate threshold as the single largest attrition point. ICU outcome data demonstrate that high-compliance episodes achieve a median LOS of 3.8 days versus 5.1 days for low-compliance episodes, with antibiotic timing showing a dose-response relationship from 2.95 days (30-60 min) to 4.74 days (*>*6 hours). Recurrent sepsis patients show only a 1.3% compliance improvement between episodes, indicating that institutional rather than individual-level interventions are required for meaningful adherence gains.

| Metric | Value | Metric | Value |
| --- | --- | --- | --- |
| *STUDY COHORT* |  | *MISSING DATA* |  |
| Total Sepsis Patients | 2,000 | R2: Antibiotic Timing | 31.6% |
| Total Sepsis Episodes | 2,438 | R3: Lactate Timing | 24.4% |
| Recurrent Sepsis (14%) | 284 | R1: Blood Culture | 8.8% |
| Unique Drug Strings | 1,691 | R5: Fluids | 6.7% |
| *NORMALIZATION* |  | *CONDITIONAL ACTIVATION* |  |
| Drug Class. Agreement | 94.26% | R4: Repeat Lactate | 912 (37%) |
| Cohen's Kappa ($\kappa$) | 0.65 | R5: Fluid Resusc. | 1,052 (43%) |
| Embedding Val. Rate | 98.59% | R6: Vasopressors | 776 (32%) |
| Agreed Antibiotics | 126 | R7: MAP Recovery | 686 (28%) |
| Agreed Vasopressors | 7 | R8: Lac. Clearance | 826 (34%) |
| *COMPLIANCE RULE ($\mu$)* |  | *ICU OUTCOMES* |  |
| R1: Blood Culture | 0.36 | Low Compliance LOS | 5.1 days |
| R2: Antibiotics | 0.24 | Med. Compliance LOS | 4.1 days |
| R3: Lactate | 0.39 | High Compliance LOS | 3.8 days |
| R4: Repeat Lactate | 0.40 | Compliance-LOS $r$ | -0.026 |
| R5: Fluid Resusc. | 0.41 | Trend (Days/1% Comp.) | -0.006 |
| R6: Vasopressors | 0.73 | Sample Size ($n$) | 2,362 |
| *TIMING & OUTCOMES* |  | *WORST-RULE IMPACT* |  |
| Meet 1-Hr Target | 13% | R2 (Antibiotic) LOS | 7.0 days |
| 30-60 min Median LOS | 2.95 d | R3 (Lactate) LOS | 5.0 days |
| 120-180 min Median LOS | 4.34 d | R1 (Blood Cult.) LOS | 4.1 days |
| 360+ min Median LOS | 4.74 d | R5 (Fluids) LOS | 2.9 days |
| *CLINICAL CASCADE* |  | *PHASE PERFORMANCE ($\mu$)* |  |
| Blood Culture Obtained | 77% | Phase 1 (Hour-1) | 0.00 |
| IV/IM Antibiotic Given | 68% | Phase 2 (Resuscitation) | 0.67 |
| Lactate Measured | 76% | Phase 3 (Response) | 1.00 |
| Fluids Administered | 36% | *RECURRENT SEPSIS* |  |
| Vasopressor Started | 37% | 1st → 2nd $\mu$ Imp. | +1.3% |
| *CASCADE DROP-OFFS* |  | *EPISODE COMPLIANCE ($\mu$)* |  |
| BC → Antibiotic | ↓23% | 1st Episode | 0.362 |
| Antibiotic → Lactate | ↓11% | 2nd Episode | 0.375 |
| Lac. → Elev. Lac. | ↓51% | 3rd+ Episodes | 0.375 |

### J. Quality of Life Implications of Sepsis Bundle Compliance

Although the primary outcomes of this study focus on ICU length of stay and temporal adherence metrics, the findings carry direct implications for sepsis survivor quality of life (QoL). Sepsis is increasingly recognized as a disease whose burden extends well beyond acute mortality, with a substantial proportion of survivors experiencing Post-Intensive Care Syndrome (PICS), characterized by cognitive impairment, psychological sequelae including PTSD and depression [29], and persistent physical disability that can endure for months to years after discharge [30]. The 1.3-day difference in median ICU length of stay between high- and low-compliance tertiles (3.8 versus 5.1 days) is not merely an administrative metric; prolonged ICU exposure is independently associated with greater muscle wasting, higher rates of delirium, increased sedation burden, and worse functional recovery trajectories, all of which are established antecedents of diminished long-term QoL. The dose-response relationship observed between antibiotic delay and ICU duration further implies that Hour-1 bundle failures compound QoL harm beyond survival: each additional hour of delayed antimicrobial therapy increases organ dysfunction exposure, raising the probability of long-term renal, pulmonary, and neurocognitive sequelae. While MIMIC-IV does not contain patient-reported outcome measures or post-discharge functional assessments, the graded compliance frame-work introduced here is architecturally compatible with QoL-linked datasets such as EHR-linked registry data or prospective follow-up cohorts. Future work integrating compliance scores with instruments such as the SF-36, EQ-5D [31], or the PROMIS physical function [32] battery would allow direct quantification of the QoL gradient attributable to bundle adherence, transforming the pipeline from a retrospective quality audit tool into a prospective predictor of survivorship outcomes.

## References

[1] Surviving sepsis campaign 2021 adult guidelines — sccm.

[2] Fang Gao, Teresa Melody, Darren F Daniels, Simon Giles, and Samantha Fox. The impact of compliance with 6-hour and 24-hour sepsis bundles on hospital mortality in patients with severe sepsis: a prospective observational study. Critical care, 9(6):R764, 2005.

[3] Je Sung You, Yoo Seok Park, Sung Phil Chung, Hye Sun Lee, Soyoung Jeon, Won Young Kim, Tae Gun Shin, You Hwan Jo, Gu Hyun Kang, Sung Hyuk Choi, et al. Relationship between time of emergency department admission and adherence to the surviving sepsis campaign bundle in patients with septic shock. Critical Care, 26(1):43, 2022.

[4] Elisa Damiani, Abele Donati, Giulia Serafini, Laura Rinaldi, Erica Adrario, Paolo Pelaia, Stefano Busani, and Massimo Girardis. Effect of performance improvement programs on compliance with sepsis bundles and mortality: a systematic review and meta-analysis of observational studies. PloS one, 10(5):e0125827, 2015.

[5] Mimic-iv v3.1.

[6] Stuart J Nelson, Kelly Zeng, John Kilbourne, Tammy Powell, and Robin Moore. Normalized names for clinical drugs: Rxnorm at 6 years. Journal of the American Medical Informatics Association, 18(4):441–448, 2011.

[7] Meicheng Yang, Hui Chen, Wenhan Hu, Massimo Mischi, Caifeng Shan, Jianqing Li, Xi Long, and Chengyu Liu. Development and validation of an interpretable conformal predictor to predict sepsis mortality risk: retrospective cohort study. Journal of Medical Internet Research, 26:e50369, 2024.

[8] Laura Evans, Andrew Rhodes, Waleed Alhazzani, Massimo Antonelli, Craig M Coopersmith, Craig French, Flávia R Machado, Lauralyn Mcintyre, Marlies Ostermann, Hallie C Prescott, et al. Surviving sepsis campaign: international guidelines for management of sepsis and septic shock 2021. Critical care medicine, 49(11):e1063–e1143, 2021.

[9] Mehak Gupta, Brennan Gallamoza, Nicolas Cutrona, Pranjal Dhakal, Raphael Poulain, and Rahmatollah Beheshti. An extensive data processing pipeline for mimic-iv. In Machine learning for health, pages 311–325. PMLR, 2022.

[10] Steven Horng, David A Sontag, Yoni Halpern, Yacine Jernite, Nathan I Shapiro, and Larry A Nathanson. Creating an automated trigger for sepsis clinical decision support at emergency department triage using machine learning. PloS one, 12(4):e0174708, 2017.

[11] Himanshu Tripathi. Experimental approach toward training and analysing siamese deep neural network for sentence with no repeated expressions. In 2023 14th International Conference on Computing Communication and Networking Technologies (ICCCNT), pages 1–5. IEEE, 2023.

[12] Bikram Pratim Bhuyan, Amar Ramdane-Cherif, Ravi Tomar, and TP Singh. Neuro-symbolic artificial intelligence: a survey. Neural Computing and Applications, 36(21):12809–12844, 2024.

[13] Mona Alshahrani, Mohammad Asif Khan, Omar Maddouri, Akira R Kinjo, Núria Queralt-Rosinach, and Robert Hoehndorf. Neuro-symbolic representation learning on biological knowledge graphs. Bioinformatics, 33(17):2723–2730, 2017.

[14] Chaoyi Wu, Xiaoman Zhang, Yanfeng Wang, Ya Zhang, and Weidi Xie. K-diag: Knowledge-enhanced disease diagnosis in radiographic imaging. arXiv preprint arXiv:2302.11557, 2023.

[15] Lauren Nicole DeLong, Ramon Fernández Mir, Zonglin Ji, Fiona Niamh Coulter Smith, and Jacques D Fleuriot. Neurosymbolic ai for reasoning on biomedical knowledge graphs. arXiv preprint arXiv:2307.08411, 2023.

[16] Oswaldo Solarte Pabón, Orlando Montenegro, Maria Torrente, Alejandro Rodríguez González, Mariano Provencio, and Ernestina Menasalvas. Negation and uncertainty detection in clinical texts written in spanish: a deep learning-based approach. PeerJ Computer Science, 8:e913, 2022.

[17] Long Chen. Attention-based deep learning system for negation and assertion detection in clinical notes. International Journal of Artificial Intelligence and Applications (IJAIA), 10(1), 2019.

[18] Yuelyu Ji, Zeshui Yu, and Yanshan Wang. Assertion detection in clinical natural language processing using large language models. In 2024 IEEE 12th International Conference on Healthcare Informatics (ICHI), pages 242–247. IEEE, 2024.

[19] Subhankar Maity and Manob Jyoti Saikia. Large language models in healthcare and medical applications: a review. Bioengineering, 12(6):631, 2025.

[20] Guangyu Wang, Guoxing Yang, Zongxin Du, Longjun Fan, and Xiaohu Li. Clinicalgpt: large language models finetuned with diverse medical data and comprehensive evaluation. arXiv preprint arXiv:2306.09968, 2023.

[21] Mahmud Omar, Vera Sorin, Jeremy D Collins, David Reich, Robert Freeman, Nicholas Gavin, Alexander Charney, Lisa Stump, Nicola Luigi Bragazzi, Girish N Nadkarni, et al. Multi-model assurance analysis showing large language models are highly vulnerable to adversarial hallucination attacks during clinical decision support. Communications Medicine, 5(1):330, 2025.

[22] Elham Asgari, Nina Montaña-Brown, Magda Dubois, Saleh Khalil, Jasmine Balloch, Joshua Au Yeung, and Dominic Pimenta. A framework to assess clinical safety and hallucination rates of llms for medical text summarisation. NPJ digital medicine, 8(1):274, 2025.

[23] Denis McInerney, Geoffrey Young, Jan-Willem van de Meent, and Byron C Wallace. Chill: Zero-shot custom interpretable feature extraction from clinical notes with large language models. In Findings of the Association for Computational Linguistics: EMNLP 2023, pages 8477–8494, 2023.

[24] Md Mehedi Hasan, Rafid Mostafiz, Md Abir Hossain, and Bikash Kumar Paul. Clin-llm: A safety-constrained hybrid framework for clinical diagnosis and treatment generation. arXiv preprint arXiv:2510.22609, 2025.

[25] Goli Arji, Hossein Ahmadi, Mehrbakhsh Nilashi, Tarik A Rashid, Omed Hassan Ahmed, Nahla Aljojo, and Azida Zainol. Fuzzy logic approach for infectious disease diagnosis: A methodical evaluation, literature and classification. Biocybernetics and biomedical engineering, 39(4):937–955, 2019.

[26] S Kumar, M Patel, BB Jayasingh, M Kumar, Z Balasm, and S Bansal. Fuzzy logic-driven intelligent system for uncertainty-aware decision support using heterogeneous data. J. Mach. Comput, 5(4), 2025.

[27] Pouria Mortezaagha and Arya Rahgozar. An auditable pipeline for fuzzy full-text screening in systematic reviews: Integrating contrastive semantic highlighting and llm judgment. arXiv preprint arXiv:2508.15822, 2025.

[28] Alan E Jones, Nathan I Shapiro, Stephen Trzeciak, Ryan C Arnold, Heather A Claremont, Jeffrey A Kline, Emergency Medicine Shock Research Network (EMShockNet) Investigators, et al. Lactate clearance vs central venous oxygen saturation as goals of early sepsis therapy: a randomized clinical trial. Jama, 303(8):739–746, 2010.

[29] Dale M Needham, Judy Davidson, Henry Cohen, Ramona O Hopkins, Craig Weinert, Hannah Wunsch, Christine Zawistowski, Anita Bemis-Dougherty, Susan C Berney, O Joseph Bienvenu, et al. Improving long-term outcomes after discharge from intensive care unit: report from a stakeholders’ conference. Critical care medicine, 40(2):502–509, 2012.

[30] Gautam Rawal, Sankalp Yadav, and Raj Kumar. Post-intensive care syndrome: an overview. Journal of translational internal medicine, 5(2):90–92, 2017.

[31] Brian H Cuthbertson, Andrew Elders, Sally Hall, Jane Taylor, Graeme MacLennan, Fiona Mackirdy, Simon J Mackenzie, Scottish Critical Care Trials Group, and the Scottish Intensive Care Society Audit Group. Mortality and quality of life in the five years after severe sepsis. Critical Care, 17(2):R70, 2013.

[32] Kewalin Pongsuwun, Wimolrat Puwarawuttipanit, Sunisa Nguantad, Benjakarn Samart, Khalinee Saikaew, and Suebsarn Ruksakulpiwat. Factor impacting quality of life among sepsis survivors during and after hospitalization: A systematic review of current empirical evidence. Journal of Multidisciplinary Healthcare, pages 3791–3802, 2024.

